# Rare copy number variation in Post-traumatic stress disorder using whole genome sequencing and biobank data

**DOI:** 10.64898/2026.09.11.26362867

**Authors:** Sydney Kramer, Saeed Farajzadeh Valilou, MadhurBain Singh, Stephen Vieno, Roseann E. Peterson, Christopher Chatzinakos, Bradley T. Webb, Jonathan R. I. Coleman, Ananda B. Amstadter, Amanda Elswick Gentry, Hermine H. Maes, Brien P. Riley, Kenneth Kendler, Christina Sheerin-Smith, Tan-Hoang Nguyen

## Abstract

Post-traumatic stress disorder (PTSD) has a significant genetic component (*h*^2^ = 24-72%). While the majority of prior studies have utilized common variants, recent research suggests rare variants also contribute to PTSD liability. Here, we examined the role of rare copy number variants (rCNVs; MAF < 1%) from multi-ancestry whole-genome sequencing data in the All of Us Research Program, using an electronic health record-defined PTSD phenotype. Genome-wide burden analyses utilizing all rCNV lengths revealed significant associations for deletion count across European-like (EUR-like; β = 0.005, SE = 0.001, *P* = 5.99 x 10^-6,^, *FDR P* = 3.59 x 10^-5^) and African-like (AFR-like; *β* = 0.003, SE = 0.001, *P* = 0.005, *FDR P* =0.02) ancestries. We conducted rCNV burden analyses within and across EUR-like (N case = 1,179, N control = 33,689), AFR-like (N case = 504, N control = 12,216), and Admixed American (AMR-like) (N case = 418, N control = 11,935) ancestries using three separate domains of gene-sets previously implicated in neuropsychiatric disorders: neurodevelopmental disorders (NDD; N = 53), abnormal behavior mouse mutant-derived (N = 146), and PTSD (N = 5). Within-ancestry analyses across all three domains yielded one significant brain-expressed gene-set within the EUR-like cohort. Meta-analyzing across ancestries identified 11 NDD gene-sets enriched for rCNV count (FDR *P* < 0.05). rCNVs with lengths greater than 10kb were examined, but observed no significant signals. Future research using larger samples and detailed PTSD phenotypes, integrated with functional genomic data and brain-region-specific expression profiles, is needed to fully map the disorder’s genetic landscape.

## Introduction

Posttraumatic stress disorder (PTSD) is a stress-related mental health disorder affecting from ∼4% to 17% of people exposed to a traumatic event^1^. Genetic factors play a significant role in PTSD risk, with heritability estimates from twin studies, *h*^2^, ranging from 24-72%^2–5^ and estimates from molecular studies of single-nucleotide polymorphisms (SNPs), *h*^2^_SNP_, ranging from 5-20% ^6–8^. Recent findings from the Psychiatric Genomics Consortium for PTSD (PGC-PTSD) have identified 95 significant genome-wide association study (GWAS) loci and 43 potentially causal genes^7^. However, focusing exclusively on common variation leaves gaps in our understanding of how rare genetic variants contribute to PTSD liability. Given PTSD is a complex disorder in which genetic susceptibility interacts with environmental stressors, identifying rare CNVs may improve our understanding of the genetic architecture of the disorder.

Despite their low population prevalence, copy-number variants often disrupt multiple genes and therefore can have larger phenotypic consequences by disrupting gene dosage or regulatory functions^9–13^. While rare variation has been understudied, available evidence suggests that PTSD is associated with rare single-nucleotide variants^14^ and rare copy-number variants (rCNVs; minor allele frequency [MAF] < 0.01)^15^. The one prior rCNV analysis conducted for PTSD used microarray data^15^ and suggested a prominent role of neurodevelopmental disorder (NDD) genes in risk for PTSD.

Here, we conducted whole-genome, gene-set, and regional burden analyses for rCNVs in PTSD using the latest whole-genome sequencing (WGS) and phenotypic data across the largest ancestry groups from the All of Us Research Program (AoU). First, we sought to replicate this prior rCNV work evaluating the association of 1) region-level, 2) genome-wide, and 3) neurodevelopmental gene-set burden with PTSD^15^. Extending on this work, we leveraged high-resolution CNVs through WGS and additional enrichment analyses to identify significant biological pathways by employing public reference atlases^7,16–18^. We conducted gene-set analyses using gene-sets commonly associated with PTSD across various genomic methods, along with gene-sets from mouse mutant studies previously implicated in neuropsychiatric disorders^7,16–18^. Significantly, we also extended all region and gene-set level analyses to AFR-like and AMR-like ancestry groups. This study was designed to evaluate two primary hypotheses: 1) analysis of large-scale WGS and multi-ancestry data would help identify rCNVs and gene-sets enriched with rCNVs associated with PTSD, and 2) meta-analyzing rCNV gene-set summary statistics across ancestries would increase the power to detect significant biological pathways associated with PTSD.

## Methods

The analysis overview is described in figure 1. A summary of the analyses is provided below, and comprehensive methodological steps are described in the Supplementary Information.

**Figure 1.**
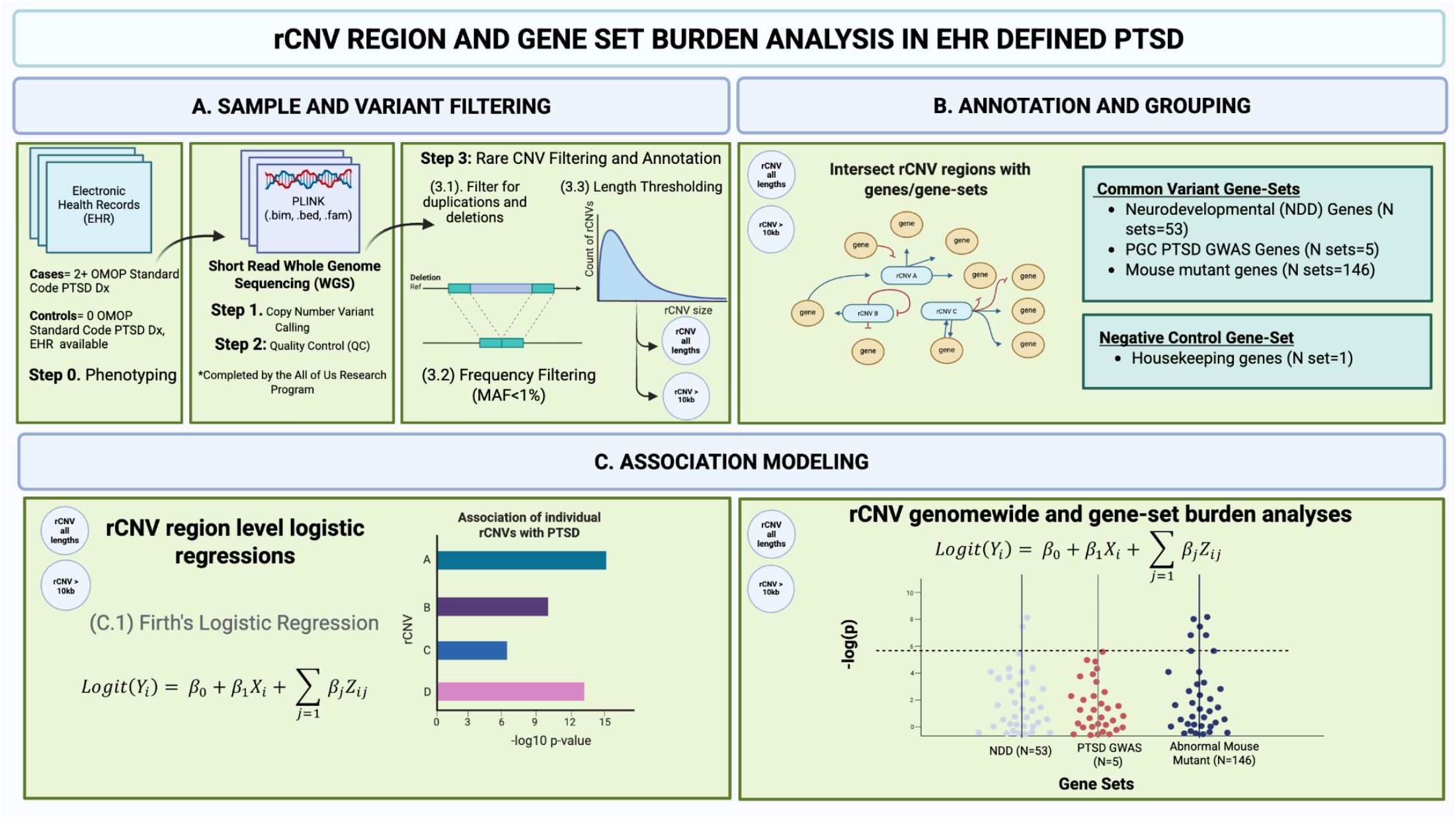
rCNV Region and Gene-Set Analysis Overview. A). OMOP = Observational Medical Outcomes Partnership (OMOP) Common Data Model (CDM). B) rCNVs were overlapped to gene-sets based on two rCNV thresholds: all rCNV lengths and rCNV’s with a total length greater than 10 kb. C). The formula is written for a single individual, *i*: *Y_i_* represents the case/control status (1 = case, 0 = control); *β*_O_ represents the intercept; *β*_1_ represents the coefficient of *X_i_*; *X_i_* represents the count of rCNVs (deletions, duplications, or total duplications and deletions within a gene-set). *β*_j_represents the corresponding coefficient for covariate *j* for individual *i*. *Z_i_*_j_ represents the value for the covariate *j* for individual *i*. The image was created with Biorender.com

### Phenotyping

#### Electronic Health Record (EHR) derived PTSD

Over 340 recruitment sites across the United States have enrolled participants with health questionnaires, EHR, physical measurements, digital health technology, and a range of genotypic data in AoU. Given the range of health care systems represented by EHR data in AoU^19^, we utilized a standardized PTSD EHR code. AoU relies on the Observational Medical Outcomes Partnership (OMOP) Common Data Model (CDM) Version 5 to standardize all EHR codes across the multiple hospital systems into a standard code. Participants were identified as cases if their medical records contained at least two entries of the PTSD standard code (436676). While prior studies have expanded EHR codes to include the broader stress disorder spectrum (e.g. adjustment disorders)^7,15^, we have restricted cases to a narrow definition of PTSD (see Athena documentation: https://athena.ohdsi.org/search-terms/terms/436676). We posited that individuals with a narrow definition of PTSD would display more severe symptom profiles and, therefore, would be more likely to be enriched for rare variants. Prior research using biobank data on major depressive disorder (MDD) has shown that narrowly defined phenotypes provide MDD-specific significant loci, whereas broadly defined MDD phenotypes yield nonspecific loci related to MDD^20^. Individuals with broader stress disorders may or may not eventually transition to a narrow definition PTSD EHR code^21^. Controls were required to have EHR records (‘has_ehr_data’) and zero occurrences of the standard code used for cases.

#### Whole-genome sequencing data

Short read WGS (srWGS) data in AoU were sequenced with the aim of a minimum coverage of 30× using Illumina NovaSeq 6000 and DRAGEN v3.7.8 software at the AoU Genome Centers (GC) and Data Research Center (DRC)^22^. AoU GC first called single samples and then jointly called variants in the WGS dataset, which comprises 414,830 individuals in the Curated Data Repository Version 8 (CDRv8) dataset. The primary sample quality control (QC) was performed by AoU (Genomic Research Data Quality).

Following the initial population stratification, a diverse global cohort was retained for analysis, consisting of 76,121 individuals of African-like (AFR-like) ancestry, 86,367 Admixed American-like (AMR-like), and 220,171 European-like (EUR-like). A total of 34,868 EUR-like, 12,720 AFR-like, and 12,353 AMR-like individuals were retained for analyses (Supplementary Methods, Supplementary Table 1).

### Variant Calling and Annotation

#### Structural Variants

CDRv8 SVs were called from CRAM files aligned with DRAGEN v3.4.12 in the CDRv7 release. Importantly, variants were not recalled during the CDRv8 release, however extra steps were taken to refine the dataset and remove samples that withdrew from v7. As a result, the SV data in v8 is aligned to DRAGEN v3.4.12, but srWGS data in CDRv8 were aligned to DRAGEN v.3.7.8. Genomic data were then processed by the GC and DRC, and a subset of srWGS samples (N = 97,061) underwent short read structural variant calling in CDRv7 with the GATK-SV pipeline^23^. GATK-SV runs multiple SV callers to increase sensitivity and leverage multiple types of evidence. Over 1.5 million SV sites were identified (https://support.researchallofus.org/hc/en-us/articles/27496716922900-All-of-Us-Short-Read-Structural-Variant-Quality-Report). While the full spectrum of high-quality SVs was called by AoU, our analyses focused on duplications and deletions to provide the most highly powered sample. The study cohort was drawn from the 97,061 individuals with high-quality SV calls generated in the AoU SV CDRv8 release, which served as the source population for our case and control selection.

#### Analytic Plan

To maximize the utility of our data, rCNV (MAF < 0.01) analyses were performed using two distinct strategies. We first evaluated all detected rCNVs to capitalize on the high resolution afforded by WGS. We then repeated the analysis restricting rCNVs to those greater than 10 kb, ensuring comparability with standard array-based findings. Prior rCNV work has relied on CNVs called from microarray data, which reliably call only CNVs greater than 10 kb. Given our use of rCNVs called from WGS, with 71% of rare autosomal CNVs below 10 kb, we conducted gene-set analyses across two thresholds: 1) rCNVs of all lengths and 2) rCNVs >= 10 kb. PLINK 2.0^24^ was utilized for rCNV analysis. Firth’s logistic regression^25^was utilized to conduct genome-wide and gene-set burden analyses in R (v.4.3.0) across unrelated participants genetically similar to European-like (EUR-like; N case = 1,179, N control = 33,689), African-like (AFR-like; N case = 504, N control = 12,216), and Admixed American-like (AMR-like; N case = 418, N control = 11,935) ancestry groups (Supplementary Methods, Supplementary Table 1).

##### Analysis of region-level CNVs

We analyzed CNV regions (duplications and deletions) with PLINK 2.0^24^ to obtain region-level p-values (Supplementary Methods). Given the high sparsity matrices due to our use of rCNVs, we utilized Firth’s logistic regression for rCNV region-level analyses. While traditional logistic regressions suffer from biased parameter estimates in the presence of small sample sizes and quasi or complete separation in the presence of rare events, Firth’s regression is able to address these issues by introducing a likelihood function based on Jeffery’s invariant prior^25^. rCNV region-level logistic regressions included the covariates: age at DNA collection, genetically inferred sex that is concordant with self-reported sex, and the first ten within-ancestry genetic principal components (PCs). Due to PLINK’s instability with minor allele counts (MAC) below 20 and given the AoU Data and Dissemination Policy, we restricted rCNVs to those with an effective carrier count greater than 20 (Supplementary Methods). For these tests, a Bonferroni correction *P* < 0.05 (*P_BF_*) was considered statistically significant, and an odds ratio of 1.2 was used as our threshold for prioritization for rCNV region analyses, based on prior rCNV work on a similar phenotype in a large-scale biobank^26^. To maintain strict control over Type I error rates across our regional rCNV analyses, multiple testing correction was applied utilizing a stringent genome-wide Bonferroni threshold (P < 0.05) rather than a False Discovery Rate (FDR) approach^28^. This choice was directly motivated by our reliance on ancestry specific allele frequencies, which exerts a significantly different statistical impact on region level locus analyses compared to aggregate gene-set analyses. While gene-set burden analyses collapse variants across broad functional pathways, inherently smoothing out localized frequency, region-level analyses evaluate tens of thousands of individual genomic regions independently. P-values, *β* coefficients, odds ratios (OR), and standard errors (SE) from PLINK 2.0 were used to infer associations between rCNVs and PTSD.

##### Analysis of CNV burden

CNV burden was examined across gene-set and genome-wide levels as conducted in a recent study^15^. Broadly, the genome-wide burden associated with PTSD was tested using Firth’s logistic regression for 1) length (mb) of duplications and deletions separately, 2) count of duplications and deletions separately, 3) total length (mb) of CNVs, and 4) total count of rCNVs. Gene and gene-set analyses calculated overlap between rCNVs on the gene and gene-set level (Supplementary Methods). For each analysis, a binary value (1 for overlap with an rCNV, 0 for no overlap) was used (Supplementary Files [Rare CNV Gene-Set Overlap Summary Statistics]). rCNVs were considered to be overlapping a gene if the rCNV overlapped the gene by at least 1 bp. Covariates included in the gene-set or genome-wide burden analyses differed between the two analyses, as described below. We conducted both genome-wide and gene-set analyses across the three largest ancestry groups (EUR-like, AFR-like, AMR-like).

#### Genome-wide burden analysis

We conducted genome-wide rCNV burden analyses for deletions and duplications, regardless of gene overlap, across count, length (megabases [Mb]), and the combined count/length using Firth’s penalized likelihood logistic regression in R (v.4.3.0). Genome-wide rCNV burden analyses were completed using 1) all rCNVs and 2) restricting rCNVs to greater than or equal to 10 kb. The primary model using Firth’s penalized likelihood logistic regression adjusted for age at DNA collection, biological sex, and the first five genetic PCs to account for population stratification^27^. The inclusion of five principal components was selected based on scree plots across ancestry groups. Five PCs were maintained in order to directly compare to prior work^15^. Similarly, we conducted sensitivity analyses using a GLM logit link function under a binomial distribution to directly compare to prior work (Supplementary Methods). Multiple testing was accounted for using the Benjamini-Hochberg false discovery rate (FDR) procedure^28^ across all genome-wide metrics, with a significance threshold of FDR *P* < 0.05.

##### Gene-set CNV burden analysis

We tested multiple gene sets derived from different omics sources (Supplementary Methods). CNVs commonly disrupt multiple genes; therefore, an underlying biological signal would be expected to manifest as a convergence of evidence across differing functional genomic analyses. Enrichment of rCNV burden using Firth’s penalized logistic regression was completed across: 1) Five PGC PTSD gene-sets^7^; 2) 53 previous NDD gene sets^15^; 3) 146 abnormal behavior gene sets derived from mouse mutants with central nervous system (CNS) phenotypes, which have been curated by neuropsychiatric disorder studies^16–18^; and 4) one negative control housekeeping gene set^29^. Our negative control gene set consisted of a large number of genes (N=197) under high evolutionary constraint which helped to provide a stable mutation resistant baseline minimizing statistical noise. Multiple testing was accounted for using the Benjamini-Hochberg false discovery rate (FDR) procedure^28^ across all gene-sets and events, with a significance threshold of FDR *P* < 0.05. We adjusted for all tested gene-sets (N=204 total gene-sets).

We controlled for average rCNV length and total rCNV count to mitigate the possibility of background confounding genomic instability within cases. Specifically, our inclusion of average rCNV length mitigates possible bias from differences in rCNV rate differing between cases and controls, while genome-wide total rCNV length adjusts for the overall rCNV burden between cases and controls^30^. Using this approach, we aimed to properly assess the association of rCNVs and gene-sets independently of overall genomic burden and rCNV size. We examined several different combinations of covariates (Formulas in Supplementary Methods). However, our primary approach followed previously established frameworks^15^ and tested the three gene set level burden metrics: deletion count, duplication count, and total rCNV count within each gene set. For each metric, we fitted Firth’s logistic regression model of the following form, adjusting for genome-wide average rCNV length, genome-wide total rCNV length, the first five within-ancestry genetic PCs, age, and sex. The inclusion of five within-ancestry PCs was chosen to remain consistent with previous large-scale rCNV work in PTSD^15^.

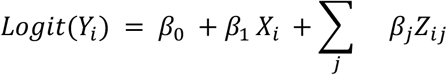

where *Y_i_* represents the case/control status of individual *i* (1 = case, 0 = control). *β*_O_ represents the intercept. *β*_1_ represents the coefficient of *X_i_*. *X_i_* represents the count of rCNVs (deletions, duplications, or total duplications and deletions within a gene-set). *β*_j_represents the corresponding coefficient for covariate *j* (e.g, age, sex) for individual *i* and *Z_i_*_j_ represents the value for the covariate *j* for individual *i*.

##### Weighted Stouffer’s Meta-Analysis

For cross-ancestry meta-analyses, we utilized a weighted Stouffer’s z-score test^31^ to determine if the cumulative pooling of z-score estimates across ancestries leads to significant gene-set associations. To ensure z-scores were not potentially canceled out by other ancestry groups, we utilized the absolute z-score to test the alternative hypothesis of whether the global pooled effect size was different from 0. By pooling absolute z-score burden estimates across EUR-like, AFR-like, and AMR-like groups^32^, we aimed to identify gene-sets associated with PTSD regardless of the direction of effect. This conservative, direction-agnostic approach was adopted to mitigate the impact of high sampling variance and potentially unreliable effect directions within the smaller AFR and AMR cohorts. Weights within each ancestry (EUR-like, AFR-like, and AMR-like) were defined as the square root of the cohort sample size (*w_i_* =3*N_i_*), ensuring that cohorts with higher statistical power contributed proportionally to the combined z-score.

## Results

### rCNV Region Analysis

We first examined the association between rCNV regions and PTSD status across all rCNV lengths (Supplementary Table 3). We observed one Bonferroni-significant rCNV region in EUR-like participants (N regions = 35,250). The most significant region was a deletion in 1p31.1 (*P_BF_* = 0.002, OR = 7.15) overlapping the *KLF18* gene, previously associated with neurodevelopmental delays^33^. Within the AFR-like cohort, we identified three significant rCNV regions (N regions = 51,954). These included a highly significant duplication mapping to the 20q13.33 cytoband (*P_BF_* = 0.002, OR = 6.16), which spans an interval of 126 bp and directly overlaps the *RBBP8NL* gene. In contrast, the remaining two loci, a deletion on cytoband 22q12.1 (*P_BF_*= 0.031, OR = 7.09) and a duplication on cytoband 13q31.1 (*P_BF_* = 0.044, OR = 4.11), both mapped to completely intergenic genomic regions. 20q13.33 has been associated with an array of developmental phenotypes and may be particularly involved in cognitive development ^34^. Notably, both the 22q12.1 and 13q31.1 regions have been previously implicated in neurodevelopmental and congenital phenotypes ^35,36^. The deletion identified at 22q12.1 maps adjacent to the well-characterized 22q11.2 microdeletion region responsible for DiGeorge syndrome^37^, a pathogenic locus strongly tied to cognitive impairment and neuropsychiatric risk. In the AMR-like cohort (N regions = 39,216), we identified a genome-wide significant regional deletion on the 4q35.2 cytoband (*P_BF_* = 0.045, OR = 8.03). This deletion spans a precise interval of 824 bp and maps to an intergenic locus, further mapping out the non-coding rCNV landscape associated with PTSD risk. 4q35.2 has been previously associated with intellectual disability and co-morbid psychiatric disorders^38^. Genomic inflation was properly controlled within the EUR-like group (EUR-like lambda_1000_ = 0.96), however deflated across AFR-like and AMR-like groups (AFR-like lambda_1000_ = 0.90, AMR-like lambda_1000_ = 0.86), likely due to the smaller sample size.

We then conducted the same analyses for rCNVs with a threshold of >= 10 kb; however, we did not observe any significant associations across any ancestral group. Considering the limited overall sample size, especially for AFR-like and AMR-like groups, we view these results as preliminary.

### Genome-wide Burden Analyses

Genome-wide burden analyses across all rCNV lengths revealed significant associations for deletion (*P* = 5.99 x 10⁻⁶, FDR *P* = 3.59 x 10^-5^, OR = 1.005, 95% CI = 1.005 – 1.101) and total count (*P* = 0.005, FDR *P* = 0.01, OR = 1.002) burden. Restricting rCNVs to 10 kb or greater, we observed a nominal enrichment of deletion length burden (*P* = 0.021, FDR *P* = 0.084, OR = 1.052). The directionality of the associations was maintained; however, a reduction in overall signal was observed compared to genome-wide burden with all rCNVs. Similarly, we observed an enrichment of deletion counts in the AFR-like groups (*P* = 0.005, FDR *P* = 0.03, OR = 1.003, 95% CI= 1.001 – 1.006). No significant associations were observed in genome-wide burden analyses in the AMR-like group (Supplementary Table 4). Expanding the inclusion of rCNVs to include those less than 10 kb, a range missed by array-based studies, but possible with WGS data. We saw consistent results with previous work^15^ (Table 1) and in our 10 kb thresholded rCNV analyses (Supplementary Results).

**Table 1.** EUR-like Genome-wide Burden Association.

| Burden Metric | Length | Beta | SE | OR | 95% CI | *P* | FDR *P* |
| --- | --- | --- | --- | --- | --- | --- | --- |
| Deletion Length (Mb) | All | 0.043<br>1 | 0.0246 | 1.044 | 0.995 – 1.096 | 0.0619 | 0.12 |
|  | >= 10<br>kb | 0.050<br>7 | 0.0234 | 1.052 | 1.005 – 1.101 | 0.021 | 0.13 |
| Deletion Count | All | 0.004<br>7 | 0.0010 | 1.0047 | 1.003 – 1.007 | 5.99 x 10<sup>-6*</sup> | 3.60 x 10<sup>-5</sup> |
|  | >= 10<br>kb | 0.005<br>4 | 0.0032 | 1.005 | 0.999 – 1.012 | 0.096 | 0.19 |
| Total Length (Mb) | All | 0.029<br>4 | 0.0173 | 1.030 | 0.996 – 1.065 | 0.0844 | 0.13 |
|  | >= 10<br>kb | 0.032<br>1 | 0.0172 | 1.033 | 0.998 – 1.068 | 0.058 | 0.17 |
| Total Count | All | 0.001<br>5 | 0.0005 | 1.0015 | 1.000 – 1.003 | 0.0051* | 0.01 |
|  | >= 10<br>kb | 0.002<br>7 | 0.0017 | 1.003 | 0.999 – 1.006 | 0.114 | 0.17 |
| Duplication Length (Mb) | All | 0.022<br>1 | 0.0234 | 1.022 | 0.977 – 1.070 | 0.338 | 0.41 |
|  | >= 10<br>kb | 0.018<br>3 | 0.0245 | 1.018 | 0.971 – 1.069 | 0.449 | 0.45 |

**Table 1.** EUR-like Genome-wide Burden Association
|  |  |  |  |  |  |  |  |
| --- | --- | --- | --- | --- | --- | --- | --- |
| Duplication Count | All | 0.0005 | 0.0008 | 1.0005 | 0.999 – 1.002 | 0.490 | 0.49 |
|  | >= 10 kb | 0.0018 | 0.0021 | 1.002 | 0.998 – 1.006 | 0.449 | 0.45 |

**Note:** PTSD cases were defined using 2+ OMOP standard codes provided by AoU. PLINK analyses excluded individuals with 3rd degree relatives or closer. All rCNVs (MAF < 0.01), regardless of length, were selected for the all lengths analysis. All rCNVs with a length greater than 10 kb were selected for >10 kb analyses. mb = Mega-bases. Covariates included EUR-like within-ancestry PC1:PC5, age at DNA collection, and sex. Beta: effect size (log-odds ratio) from PLINK, SE=standard error, OR=odds ratio.

#### Gene-set burden analysis

##### NDD gene-sets

We examined rCNV burden in 53 NDD gene-sets across multiple metrics and covariates. We observed one significant gene set related to developing brain expression. Specifically, the gene-set, BspanML_lg2rpkm0.93, was enriched for duplication count (*P* = 1.4 x 10^-5^, FDR *P* = 0.008, OR = 0.97). No significant association was observed between any rCNV metric and our negative-control housekeeping gene set after multiple-testing correction (FDR *P* < 0.05 for all models). Sensitivity analyses across AFR-like and AMR-like groups did not identify any significant gene-sets across negative control and NDD gene sets. We did not observe any significant associations across any ancestral group utilizing rCNVs greater than 10 kb (Supplementary Table 5).

##### PTSD Common Variant Gene-Sets

Burden analyses using five common variant gene sets from a large-scale PGC PTSD GWAS^7^ were conducted. While we did not have any significant or nominally significant rCNV burden across any metric, we observed a notable genomic deflation in both EUR-like and AFR-like ancestry groups (EUR-like median lambda = 0.66; AFR-like lambda = 0.34), while the AMR-like group showed genomic inflation (median lambda = 1.75). However, given that each ancestry specific median lambda estimate was drawn from 15 tests, these estimates are highly sensitive to individual outlier pathways. In both EUR-like and AMR-like ancestries, the most significantly enriched gene set was a gene set identified via single cell enrichment testing in the dorsolateral prefrontal cortex. In both ancestral groups, this top gene-set was enriched for deletion count. Burden analyses using rCNVs greater than 10 kb similarly did not lead to significant or nominally significant associations across any metric (Supplementary Results, Supplementary Table 5).

##### Abnormal Behavior Gene-Sets

We examined the rCNV burden of 146 gene sets curated by previous studies for neuropsychiatric disorders^16–18^. After applying FDR correction, 1 gene-set enriched for deletion count in the EUR-like group was identified (abnormal_nervous_system_morphology; *P* = 9.2x10^-5^, FDR *P* =0.03, OR = 1.02), along with 71 nominal associations in 50 gene-sets. Similarly, one gene set, Chrna7, was enriched for genome-wide total count (*P* = 7x10^-5^, FDR *P* = 0.04) in the AMR-like group. No significant gene-sets were observed in the AFR-like ancestry group (see Supplementary Table 5). Genomic inflation (lambda_1%_ = 1.61) was evident in EUR-like, but not AFR-like (lambda_1%_ = 0.68) or AMR-like (lambda_1%_ = 1.01). We observed one gene-sets nominally significant in EUR-like and AMR-like (mir137) and seven shared in two or more ancestry groups (Supplementary Table 5).

#### Weighted Stouffer’s Meta-Analysis

##### NDD gene-sets

We meta-analyzed across all rCNV lengths. In the meta-analysis, we observed the same gene-set enriched for deletion count as in the EUR-like ancestry group (BspanML_lg2rpkm0.93; *P* meta = 9.0 x 10^-6^, FDR *P* = 0.005). This gene-set along with six additional significant gene sets were enriched for deletion and duplication count burden involved in brain expression (e.g. scRNA_Expressed_PgG2M, BspanHM_lg2rpkm3.21), synaptic functioning (e.g. PSD_BayesGrant_fullset), and constraint (e.g. gnomAD_oe_lof_upper_0.35). The remaining gene-sets were identified through mouse models and implicated cardiac muscle (PhMm_Aggr_CardvascMuscle_all) and adipose tissue defects (e.g. PhMm_Aggr_IntegAdipPigm_all). Four gene-sets enriched for deletion count were also identified in previous work^15^.We identified 35 gene-sets both nominally significant and directionally consistent across ancestries. No nominal or FDR significant gene-sets displayed significant heterogeneity (*I*^2^ > 50%) across ancestries. Following this, we meta-analyzed across rCNV lengths greater than 10 kb for NDD gene sets and did not observe any significant gene sets; however, ten nominally significant associations in eight unique gene-sets were identified. Statistical power improved for the top gene set in the EUR-like analyses (scRNA-DEGs-ExDp2; *P* meta = 0.006, Meta Z = 2.75) (Supplementary Table 6).

##### PGC PTSD gene-set

Across both rCNV length thresholds for PTSD GWAS gene-sets, we did not observe any significant gene-sets. However, nominal associations were observed across genes in the eQTL Summary-based Mendelian Randomization (SMR) analysis^7^ (see Nievergelt et al., 2024; Supplementary Table 17) of single-cell types within the dorsolateral prefrontal cortex for deletion count (*P* meta = 0.02, FDR *P* meta = 0.23, Z meta = 2.75). Although this association did not survive multiple testing correction, it points to localized rCNV burden within genes mapping to cortical cell-types implicated in the transcriptomic stress response of PTSD. Meta-analysis of rCNV regions greater than 10 kb did not show any significant gene-sets, although we observed the same gene set nominally significant in the all rCNV length analyses enriched for deletion count (*P* meta = 0.02, FDR *P* meta = 0.25, Z meta = 2.75). Notably, this gene-set was one of two gene-sets showing the same direction of effect across ancestries. Similarly, no significant heterogeneity (*I*^2^ < 50%) was observed across the nominally significant gene-set (Supplementary Table 6).

##### Abnormal behavior gene sets derived from mouse mutants with central nervous system (CNS) phenotypes

Four significant gene-sets related to associative learning (eg., abnormal_associative_learning) and constraint (eg., pLI09, essential_gene, PSD_human_core) were enriched for deletion count. 40 gene-sets showed nominal rCNV enrichment. Further, 34 out of 50 nominally significant associations shared the same effect direction across EUR-like, AFR-like, and AMR-like ancestry groups, suggesting congruence in genetic effects. The top nominal gene-sets were broadly implicated in abnormal learning and memory (e.g., abnormal learning_memory or conditioning, abnormal cued conditioning behavior), synaptic regulation (e.g., synaptome, mir137), and central nervous system morphology (e.g., abnormal hypothalamus morphology, and abnormal nervous system morphology). When restricting analysis to rCNVs > 10 kb, no gene-sets were statistically significant, though 12 nominal associations were observed. Among the top 15 associations identified in the all rCNV lengths framework, only three gene-sets remained nominally significant (*P*_meta_ < 0.05) when restricted to the 10kb threshold. No high heterogeneity (*I*^2^ > 50%) was observed in nominally significant gene-sets when restricted to lengths greater than 10 kb (Supplementary Table 6).

## Discussion

The current study analyzed rCNV called from WGS data from a multi-ancestry population biobank dataset to investigate the contribution of rCNVs to PTSD. First, we sought to examine whether the analysis of large-scale WGS and multi-ancestry data would help identify individual rCNVs and associated gene-sets. Secondly, we tested whether meta-analyzing rCNV gene-set summary statistics across ancestries would increase statistical power to detect significant biological pathways associated with PTSD. Our results provide answers to both aims. Regarding our first aim, using WGS data allowed us to detect significant rCNVs associations, genome-wide burden signals (Table 1, Supplementary Table 4) and enrichments within biologically relevant gene-sets (Table 2, Supplementary Table 5), highlighting genetic regions previously implicated in NDD and psychiatric disorders. These results underscore the potential utility of sequencing-based approaches for detecting PTSD-associated rCNVs that may have not been captured by earlier array-based studies. Regarding our second aim, multi-ancestry meta analysis increased our statistical power and uncovered a suggestive convergence of six distinct biological pathways mapping to genomic constraint, brain expression, synaptic architecture and NDD-related regions (Supplementary Table 6). These findings suggest that combining WGS-derived rCNVs with cross-ancestry analytical approaches can improve the detection of shared biological mechanisms underlying PTSD.

**Table 2.**
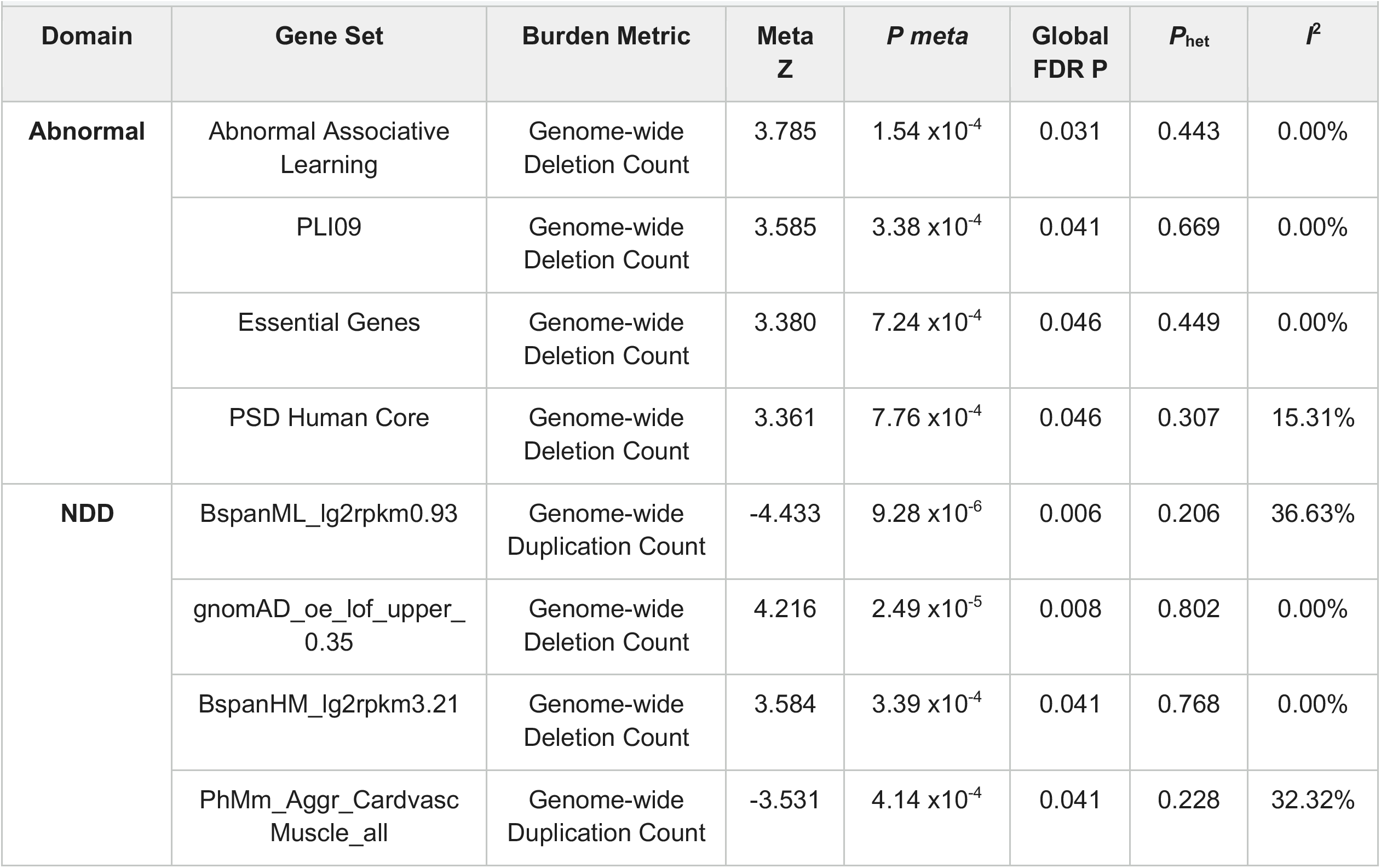
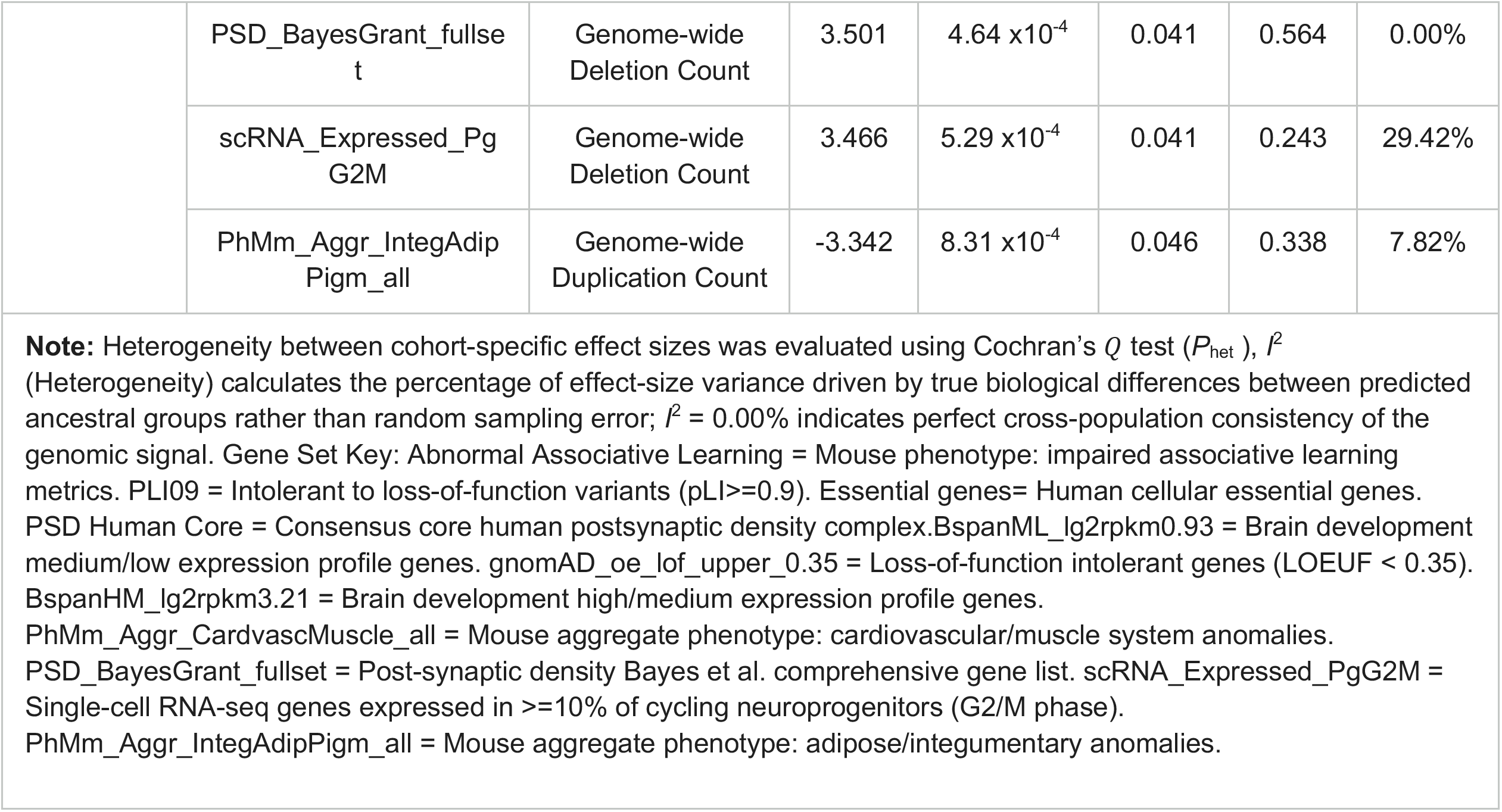
Gene-Set Meta-Analysis (EUR-like, AFR-like, AMR-like)

| Table 2. Gene-Set Meta-Analysis (EUR-like, AFR-like, AMR-like) |  |  |  |  |  |  |  |
| --- | --- | --- | --- | --- | --- | --- | --- |
| Domain | Gene Set | Burden Metric | Meta Z | $P_{\text{meta}}$ | Global FDR P | $P_{\text{het}}$ | $I^2$ |
| Abnormal | Abnormal Associative Learning | Genome-wide Deletion Count | 3.785 | $1.54 \times 10^{-4}$ | 0.031 | 0.443 | 0.00% |
| | PLI09 | Genome-wide Deletion Count | 3.585 | $3.38 \times 10^{-4}$ | 0.041 | 0.669 | 0.00% |
| | Essential Genes | Genome-wide Deletion Count | 3.380 | $7.24 \times 10^{-4}$ | 0.046 | 0.449 | 0.00% |
| | PSD Human Core | Genome-wide Deletion Count | 3.361 | $7.76 \times 10^{-4}$ | 0.046 | 0.307 | 15.31% |
| NDD | BspanML_lg2rpkm0.93 | Genome-wide Duplication Count | -4.433 | $9.28 \times 10^{-6}$ | 0.006 | 0.206 | 36.63% |
| | gnomAD_oe_lof_upper_0.35 | Genome-wide Deletion Count | 4.216 | $2.49 \times 10^{-5}$ | 0.008 | 0.802 | 0.00% |
| | BspanHM_lg2rpkm3.21 | Genome-wide Deletion Count | 3.584 | $3.39 \times 10^{-4}$ | 0.041 | 0.768 | 0.00% |
| | PhMm_Aggr_Cardvasc Muscle_all | Genome-wide Duplication Count | -3.531 | $4.14 \times 10^{-4}$ | 0.041 | 0.228 | 32.32% |
|  | PSD_BayesGrant_fullset | Genome-wide Deletion Count | 3.501 | 4.64 x10<sup>-4</sup> | 0.041 | 0.564 | 0.00% |
|  | scRNA_Expressed_PgG2M | Genome-wide Deletion Count | 3.466 | 5.29 x10<sup>-4</sup> | 0.041 | 0.243 | 29.42% |
|  | PhMm_Aggr_IntegAdipPigm_all | Genome-wide Duplication Count | -3.342 | 8.31 x10<sup>-4</sup> | 0.046 | 0.338 | 7.82% |
| **Note:** Heterogeneity between cohort-specific effect sizes was evaluated using Cochran's $Q$ test ($P_{het}$), $I^2$ (Heterogeneity) calculates the percentage of effect-size variance driven by true biological differences between predicted ancestral groups rather than random sampling error; $I^2 = 0.00\%$ indicates perfect cross-population consistency of the genomic signal. Gene Set Key: Abnormal Associative Learning = Mouse phenotype: impaired associative learning metrics. PLI09 = Intolerant to loss-of-function variants ($pLI \geq 0.9$). Essential genes= Human cellular essential genes. PSD Human Core = Consensus core human postsynaptic density complex. BspanML_Ig2rpkm0.93 = Brain development medium/low expression profile genes. gnomAD_oe_lof_upper_0.35 = Loss-of-function intolerant genes ($LOEUF < 0.35$). BspanHM_Ig2rpkm3.21 = Brain development high/medium expression profile genes. PhMm_Aggr_CardvascMuscle_all = Mouse aggregate phenotype: cardiovascular/muscle system anomalies. PSD_BayesGrant_fullset = Post-synaptic density Bayes et al. comprehensive gene list. scRNA_Expressed_PgG2M = Single-cell RNA-seq genes expressed in $\geq 10\%$ of cycling neuroprogenitors (G2/M phase). PhMm_Aggr_IntegAdipPigm_all = Mouse aggregate phenotype: adipose/integumentary anomalies. |  |  |  |  |  |  |  |

In the EUR-like cohort, we observed a significant regional deletion overlapping the *KLF18* gene, previously linked to neurodevelopmental delays and autism spectrum disorder^39,40^. Our significant findings in the AFR-like and AMR-like cohorts point toward suggestive risk loci within established neurodevelopmental bands, including a gene disruption at 20q13.33 (*RBBP8NL*) and a regional deletion adjacent to the DiGeorge syndromic region. Although several rare CNVs did not reach statistical significance, they localize to highly relevant neuropsychiatric regions, including the *NR3C1* locus (5q31-32) linked to HPA-axis regulation and PTSD pathology^41,42^ and the 1p35 neuroticism-associated locus^43^ harboring candidate anxiety genes^44^ (Supplementary Table 3). Further, these non-significant rCNV regions intersect established pleiotropic risk regions like 16p12.1, a locus heavily implicated in severe psychiatric morbidity, depression, and broader genetic vulnerability in both clinical cohorts and the general population^45–47^. This suggests a convergence in common and rare variant risk for PTSD.

Genome-wide burden results in EUR-like participants were consistent with prior work^15^ (Table 1, Supplementary Table 4) and in our 10 kb thresholded rCNV analyses. Specifically, an enrichment of deletion burden length (Mb) across both rCNV thresholds was observed in EUR-like participants. Genome-wide deletion count was significantly enriched in EUR-like and AFR-like PTSD cases using all rCNV lengths. While an increased burden of deletion count was observed in EUR-like and AFR-like PTSD cases, no significant associations with rCNV burden were observed in AMR-like participants. However, the robustness of AFR-like and AMR-like results are unclear as the current analyses are an extension of prior work which was restricted to EUR-like individuals.

Burden analyses across the three domains of gene-sets similarly showed increased significance when including all rCNV lengths and across rCNV count metrics (Supplementary Table 5 and 6). While we conducted additional sensitivity analyses using variations of covariates, we reported gene-set results using a validated framework^15^. Nevertheless, we acknowledge a need for future analyses to address the unique qualities of rCNVs called from high-resolution WGS data. There was overlap in one EUR-like gene-set (scRNA_Expressed_PgG2M) using this prior work’s covariates of genome-wide average rCNV length (Mb), genome-wide rCNV count, and the first five genetic principal components^15^. In contrast, meta-analysis across ancestries identified four overlapping gene-sets. Interestingly, these gene-sets (scRNA_Expressed_PgG2M, BspanHM_lg2rpkm3.21, PSD_BayesGrant_fullset, gnomAD_oe_lof_upper_0.35) are associated with genomic constraint, brain expression and synaptic architecture^48,49^. Finally, NDD gene-set burden analyses across all rCNV lengths in EUR-like participants identified one gene-set, involved in developing brain expression genes, enriched for duplication count.

Current literature on PTSD has largely examined common and rare genetic variation separately; however, integrating these approaches could enhance statistical power to identify genes or loci linked to this disorder. To date, large-scale sequencing data studies integrating these types of variants have not been conducted for PTSD. As a preliminary step for future analyses integrating common and rare variants, we examined rCNV burden in gene sets inferred from common variants and omics data. Our goal was to determine whether rare CNVs are significantly enriched within biological pathways already associated with PTSD from large scale common variant studies^7^. However, we did not observe any significant enrichment with the functional gene sets prioritized from these PTSD common variant studies. It is possible that the selected PTSD gene sets did not overlap with rCNV regions to the same degree as the NDD and mouse-mutant-derived abnormal behavior gene sets. For example, the median number of rCNVs (all lengths) overlapping the five PTSD gene sets was 3,661bp, while the median number of rCNVs across all gene-sets was 8,490bp. This illustrates a known limitation of gene-set burden analyses^30^, namely that large gene sets (in total number of genes contained in the gene set or the physical size of these gene sets) as well as increased sample size (in each ancestry group) would increase the probability of significant rCNV enrichment due to the cumulative physical scale of the gene-set. However, we observed that rCNVs 10kb or greater are the primary contributors to the signal within the PTSD common variant gene sets suggesting that larger structural variants may be more relevant to PTSD-associated loci, even though our current sample size lacks the statistical power to reach significance. Future studies with larger, and ancestry-diverse cohorts and expanded WGS data will be essential to clarify whether this size-dependent enrichment pattern reflects a true biological signal (Supplementary Table 6).

While the EUR-like within ancestry analysis of abnormal mouse mutant gene-sets identified only one significant gene-set enriched for deletion count (specially involved in abnormal nervous system morphology), the subsequent meta-analysis revealed four gene-sets enriched for count. Importantly, all significant gene-sets shared the same direction of effect across ancestral backgrounds. This suggests that gene sets implicated in brain morphology and neuron differentiation may have meaningful biological signals, however the current sample may not be well powered enough to identify associations. While these abnormal mouse mutant datasets provide preliminary evidence of altered neuroplasticity, this hypothesis is more reinforced when examining the NDD gene-sets. Notably, zero prioritized gene-sets with FDR significant associations demonstrated significant heterogeneity across ancestries. Additionally, multiple top nominal hits from the size-restricted (>10 kb) NDD analysis, including BspanML_lg2rpkm0.93, scRNA_Expressed_PgG2M, and PhMm_Aggr_IntegAdipPigm_all, directly overlapped with the top ten associations from the all rCNV length framework (Supplementary Table 5). This suggests that the signals identified in the all length rCNV analyses are not sequencing artifacts, but rather reflect a conserved genetic signal that is shared across ancestral backgrounds. Instead, they reflect genetic signal whose statistical power is amplified (e.g., scRNA_Expressed_PgG2M dropping from nominal *P* = 5.93 x 10^-3^ to *P* = 5.29 x 10^-4^) when short-read WGS captures the full spectrum of variants (<10 kb) previously obscured by traditional arrays.

Although much of our genome-wide burden results were in line with prior work^15^, some discrepancies are worth noting. Given our significant increase in rCNVs called from WGS, we used all rCNVs, regardless of gene overlap, rather than filtering for those overlapping genes as in prior array-based rCNV work. We hypothesize that this may be a contributing factor in the lack of congruence between our results and the previous results. Additionally, results may diverge from prior work due to MAF calculations. We utilized strict, ancestry-specific MAF filtering fit for AoU’s distinct population, whereas it is unclear in prior work if the MAF threshold for rCNVs was ancestry-specific. Across burden analyses, we observed that rCNV count emerged as the primary burden metric associated with PTSD status. This suggests that, when using sequencing-based rCNVs, the number of rare copy number variant events, rather than cumulative size, is a primary driver of association with abnormal phenotypes, particularly across neurodevelopmental phenotypes, abnormal mouse models, and PTSD-implicated functional gene-sets.

### Limitations

While we were able to utilize rCNVs called from WGS, there currently is no gold standard to QC these rCNVs. In addition, historically it has been difficult to classify SVs, as they tend to reside within repetitive DNA, which makes their characterization more difficult^50^. This is particularly relevant, given that intergenic rCNVs comprise approximately 40% of our dataset. We predict as CNV calling from WGS becomes more widespread, more refined QC pipelines will allow for increased reliability in CNV calls. Given that 71% of all rCNVs identified in the current analyses are less than 10kb and 18% are 100bp or less, it remains difficult to determine the true validation rate of these rCNV events. As most significant rCNV region-level results were small rCNVs, the current findings should be viewed as preliminary. Additionally, a large portion of significant gene-set associations were driven by rCNV count rather than total rCNV burden. Future analyses should focus on the examination of proper covariates when conducting gene-set analyses using CNVs called from WGS. Given the significant distribution in CNV size and possibility of false positive CNVs, simulations and examination of other covariates is needed. Furthermore, while analyzed across ancestral diverse populations, we limited our analyses to AoU. Future studies should validate the current results within larger biobanks.

Nuances and limitations regarding phenotyping, relevant to this and all biobank work, should be noted. Defining cases and controls using EHR data is inherently prone to bias. Future studies incorporating broader PTSD definitions, empirical examination of the genetic relationship between PTSD and adjustment disorders, and alternatively, more nuanced PTSD definitions, may help elucidate genetic signals specific to various clinical subgroups.

## Conclusion

Despite these limitations, present study findings represent the first multi-ancestry investigation of rare copy number variants in PTSD utilizing whole genome sequencing. Our analysis identified novel regions/gene-sets with biological relevance to PTSD and demonstrates alignment with common variant approaches.

## Code availability

The code for analysis could be found at https://github.com/saeedfv/AoU-CNV-Pipeline

## Author contributions

SK, SFV, SV, MS, REP, JC, ABA, CS, and THN all contributed to the study design, with THN leading the study design and formation. Phenotype curation was completed by SK, JC, ABA, HM, and CS. SK, CS, and THN wrote the manuscript draft. Analyses were done by SK. SFV, SK, and SV developed the analysis pipeline. REP, CC, MS, BW all contributed to creation of ancestral principal components. All authors provided edits to the manuscript.

## Acknowledgements

We gratefully acknowledge *All of Us* participants for their contributions, without whom this research would not have been possible. We also thank the National Institutes of Health’s *All of Us* Research Program for making available the participant data examined in this study.

The All of Us Research Program is supported by the National Institutes of Health, Office of the Director: Regional Medical Centers: 1 OT2 OD026549; 1 OT2 OD026554; 1 OT2 OD026557; 1 OT2 OD026556; 1 OT2 OD026550; 1 OT2 OD 026552; 1 OT2 OD026553; 1 OT2 OD026548; 1 OT2 OD026551; 1 OT2 OD026555; IAA #: AOD 16037; Federally Qualified Health Centers: HHSN 263201600085U; Data and Research Center: 5 U2C OD023196; Biobank: 1 U24 OD023121; The Participant Center: U24 OD023176; Participant Technology Systems Center: 1 U24 OD023163; Communications and Engagement: 3 OT2 OD023205; 3 OT2 OD023206; and Community Partners: 1 OT2 OD025277; 3 OT2 OD025315; 1 OT2 OD025337; 1 OT2 OD025276. In addition, the All of Us Research Program would not be possible without the partnership of its participants.

## Funding

5T32MH020030-25

R21MH137508

K25AA030072

## Data Availability Statement

The individual-level data underlying the findings of this study are protected under technical, legal, and policy safeguards by the National Institutes of Health (NIH) *All of Us* Research Program. To protect participant privacy and prevent re-identification, individual-level data cannot be made publicly available or distributed outside the platform. The complete, curated datasets, data dictionaries, and analytical tools used in this study are available to registered and authorized researchers within the secure, cloud-based *All of Us* Researcher Workbench. Access requirements, registration procedures, and platform data use policies can be found at https://www.researchallofus.org

## Supplemental Methods

### 1.0 Whole-genome sequencing data

Short-read whole-genome sequencing (srWGS) data in All of Us (AoU) were generated on the Illumina NovaSeq 6000 platform (30× target coverage) and processed using DRAGEN v3.7.8^51^. Joint variant calling across 414,830 participants in Curated Data Repository Version 8 (CDRv8) was performed by the AoU Genome Centers and Data Research Center, with primary genomic quality control conducted by AoU Genomic Research Data Quality. ([Genomic Research Data Quality](https://support.researchallofus.org/hc/en-us/article_attachments/40491583732116)). Additionally, samples were flagged by AoU based on three sample-level QC metrics: number of SNPs: < 2.4M and > 5.0M, number of variants not present in gnomAD ([gnomAD 3.1](https://gnomad.broadinstitute.org/news/2020-10-gnomad-v3-1-new-content-methods-annotations-and-data-availability/#sample-qc-metric-outlier-filtering)) 3.1: >100k, heterozygous to homozygous ratio (SNPs and Indel separately): > 3.3 (https://support.researchallofus.org/hc/en-us/articles/29475233432212-Controlled-CDR-Directory). As of August 2025, samples with low coverage (< 20x) were erroneously included in the WGS filesets.

Genetically inferred sex was called from array data using GenCall tool v3.0.0 ([Illuminia](https://www.illumina.com/Documents/products/technotes/technote_gencall_data_analysis_software.pdf)) and Picard 2.26.0 ([Picard](https://gatk.broadinstitute.org/hc/en-us/articles/360037593871-CollectArraysVariantCallingMetrics-Picard)). Those with discordant sex and those that reported their sex as “Other” were removed by AoU. Following this, we removed participants who reported “Intersex,” “Prefer not to answer,” “None of these fully describe me,” or who skipped the question (N < 30). In addition, DRAGEN-inferred sex was compared with self-reported sex assigned at birth for confirmation of sex concordance. Only samples with concordant male or female classifications were retained, while samples with sex mismatches or abnormal DRAGEN ploidy were excluded from the study. We manually confirmed mean coverage (threshold >= 90% at 20x). In addition, we removed samples included in the known issues with data quality issues or single variant sites missing (https://support.researchallofus.org/hc/en-us/articles/29390274413716-All-of-Us-Genomic-Quality-Report) and removed outliers that deviated from eight standard deviations (within ancestry) from the mean on key quality control metrics including: number of deletions, number of insertions, number of SNVs, number of variants not present in gnomAD 3.1 (singletons), insertion-to-deletion ratio, transition-to-transversion ratio, SNV heterozygous-to-homozygous ratio, and indel heterozygous-to-homozygous ratio^52^.

### 1.1 Genetic Similarity-Based Ancestry Assignment

Genetically inferred categorical ancestry assignment in AoU was previously completed using a newly developed pipeline *POP-MaD* (population grouping by Mahalanobis distance) pipeline^53^, using the 1000 Genomes Project Phase-3 (KGP) and the Human Genome Diversity Project (HGDP) reference panels^54^. Samples passing microarray quality control^53^were projected onto the KGP and HGDP principal components using smartPCA^55^. AoU samples were assigned to seven global populations based on genetic similarity (African/AFR, Admixed American/AMR, Central and South Asian/CSA, East Asian/EAS, European/EUR, Middle-Eastern/MID, and Oceania/OCE), defined using the minimum Mahalanobis distance from the reference populations in genetic principal components analyses (PCA). Those with a Mahalanobis distance more than 3 standard deviations from the reference group’s median Mahalanobis distance were considered ancestry group outliers. Relatedness was estimated within each assigned ancestry group using KING -related --degree 3 ^56^. The unrelated subset was defined as those with pairwise relatedness less than 3rd degree with KING (i.e., kinship coefficient < 0.5^9/2^ corresponding to 0.04419). Individuals unassigned to a global population (i.e., ancestry group outliers) and related individuals (≥ 3rd degree) were excluded from analyses. Within-ancestry genetic PCs were computed using FlashPCA2. Unrelated inliers were used for each global population analysis. This resulted in 82,675 samples assigned to the African-like (AFR-like), 94,245 assigned to Admixed American-like (AMR-like), 7,972 assigned to Central/South Asian-like (CSA-like), 10,009 to East Asian-like (EAS-like), 239,578 to European-like (EUR-like), 2,687 to Middle Eastern-like (MID-like), and 60 to Oceanian-like (OCE-like)^53^.

### 1.2 Analysis of region-level CNVs using PLINK

We analyzed CNV regions (duplications and deletions) with PLINK 2.0 to obtain region-level p-values. Low MAC can inflate the lambda values and lead to spurious associations in cases where one or a few cases have a rCNV, but the rCNV is not observed in the controls. In addition, PLINK reports unstable association metrics of MAC < 20. For these tests, we restricted rCNVs to MAC > 20 for rCNV region analyses, as testing CNVs with a MAC < 20 can inflate lambda values and lead to spurious associations.

In addition, to comply with All of Us data privacy regulations regarding small cell sizes, we implemented a rigorous downstream filtering pipeline to guarantee that no reported results were derived from ultra-rare events present in 20 or fewer individuals. While initial variant filtering utilized a standard Minor Allele Count threshold (PLINK2 --mac 21), it is possible raw allele counts can overestimate true sample sizes for structural variants due to multi-copy duplications or homozygous states. Therefore, we explicitly calculated the effective carrier count for each regional window by scaling the non-missing sample observation count (OBS_CT) by twice the variant frequency (2 * OBS_CT * A1_FREQ). Loci were strictly pruned to retain only regions backed by an effective sample size of greater than 20 true carriers, ensuring absolute compliance with the *All of Us* Research Program Data and Statistics Dissemination Policy (https://support.researchallofus.org/hc/en-us/articles/360043016291-How-to-comply-with-the-All-of-Us-Data-and-Statistics-Dissemination-Policy).

Analyses used PLINK “Unrelated Inliers” from each *POP-MaD* ancestry grouping, as PLINK is unable to account for relatedness. Duplications and deletions with a MAF < 0.01 and MAC > 20 were filtered for all regional-level analyses. MAF was determined based on VCF reported rCNV allele frequencies across each ancestry. Firth’s regression analysis in PLINK was performed to obtain p values. P-values passing False Discovery Rate (FDR) were utilized to draw inferences.

To maintain strict control over Type I error rates across our regional rCNV analyses, multiple testing correction was applied utilizing a stringent genome-wide Bonferroni threshold (P < 0.05) rather than a False Discovery Rate (FDR) approach^28^. This choice was directly motivated by our reliance on ancestry specific allele frequencies, which exerts a significantly different statistical impact on region level locus analyses compared to aggregate gene-set analyses. While gene-set burden analyses collapse variants across broad functional pathways, inherently smoothing out localized frequency, region-level analyses evaluate tens of thousands of individual genomic regions independently. Because the genetically diverse African-like ancestry (AFR-like) cohort yields a vast, dense distribution of localized, population-specific ultra-rare variants, an FDR threshold can easily suffer from background p-value inflation in a region-level matrix. Enforcing a strict Bonferroni correction controls the overall Family-Wise Error Rate (FWER), effectively protecting our rCNV region discovery analyses from the false positives generated by extreme background genetic diversity while ensuring that only highly penetrant, site-specific risk signals are declared significant.

To assess systemic genomic inflation while accounting for imbalances in cases and controls, we calculated a standardized lambda_1000_ value^57,58^. This is defined as the ratio between the median of the observed chi square association test statistic to its counterpart under the null hypothesis of no association. In this case, a value greater than 1 signals inflation in test statistics^57,58^.

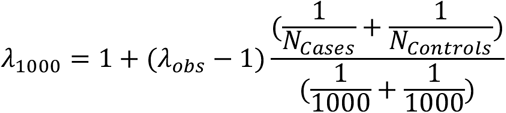

### 1.3 Analysis of CNV burden

CNVs were mapped to gene names as follows. CNV coordinate information from VCF files was converted to BED format using bcftools ^59^query. For investigation of large-scale CNVs, we intersected CNVs with cytoband information from UCSC (https://hgdownload.soe.ucsc.edu/goldenPath/hg38/database). If a single CNV overlapped multiple cytobands, their names were consolidated into a single, semicolon-separated unique label for that CNV. Following this, CNV coordinates were mapped to the genes they overlap in GENCODE gene annotation reference (<u>Release 46, GRCh38.p14)</u> using bedtools^60^. Within AoU, most autosomal rCNVs are small with 71% spanning less than 10,000 bp. CNVs may also overlap with multiple genes; therefore, each CNV was mapped to all the genes it overlaps by at least one base pair. We aggregated gene names from all individual transcript overlaps to create a consolidated list of gene names. All primary quality control filtering and statistical analyses were performed using PLINK 2.0. However, due to the deprecation of legacy recoding functionalities in the PLINK 2.0 architecture, PLINK 1.9 was specifically utilized to convert the filtered datasets and extract exact allele/variant counts. Specifically, the `--recode` function in PLINK 1.9 was used to output the necessary additive dosage and count formats required for downstream gene-set burden modeling.

#### 1.3.1 Genome wide burden analysis

We conducted genome-wide rCNV (with > 10 kb) burden analyses for deletions and duplications, regardless of gene overlap, across count, length (megabases [Mb]), and the combined count/length using Firth’s regression in R (v.4.3.0). We also report generalized linear mixed model (GLM) logistic results for PGC related models in EUR-like as a sensitivity analysis. For comparison, we also report Bonferroni-adjusted p-values, representing the most conservative threshold for family-wise error rate control. Specifically, we applied a Bonferroni correction for ten independent tests (5 burden metrics: Deletion/Duplication/Total Mb Burden and Deletion/Duplication Count; across 2 size resolutions: all rare CNVs and rCNVs > 10 kb), resulting in a p value threshold of 0.005.

#### 1.3.2 Gene-set Burden Analyses

Previous large scale genomic studies have demonstrated an enrichment of genes associated with PTSD in various brain regions (e.g., hippocampus, amygdala, and pituitary gland) which play a role across memory consolidation, emotional processing, and neuroendocrine regulation of stress response^7^. This enrichment of genes has been triangulated with functional methods using gene expression (eQTLs)^7,61^, protein abundance (pQTLs)^62^, and post-transcriptional regulation (mQTLs)^63^. We examined gene-set burden of rCNVs across gene-sets from differing functional studies which converge on common biological pathways.

We examined various gene-set models containing different covariate specifications. We wanted to include genome-wide rCNV count to ensure that the gene-set signal is independent of global burden^30^. In addition to gene-set analyses using the same covariates (top 5 PCs, average rCNV length, total rCNV count) and varying levels of predictors (total count, duplication count, deletion count) as in previous research^15^(Formula 1). We added genome-wide total rCNV count to determine if the observed gene-set associations were independent of global rCNV events. Previous research has demonstrated that GLM can inflate p-values due to sparsity in rare variant association testing^27^, however prior research has often relied on GLM for rare variant burden testing.

Gene-set burden analyses evaluated the association between rCNV burden and PTSD diagnosis across cases and controls in the following formula:

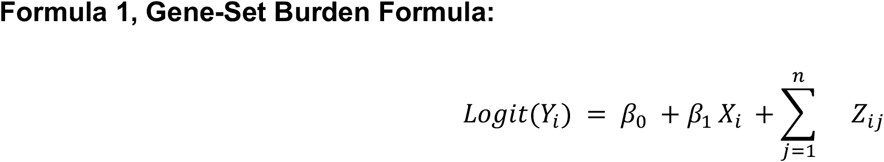

where *Y_i_* represents the case/control status of individual *i* (1 = case, 0 = control). *B*_O_ represents the intercept. *B*_1_ represents the coefficient of *X_i_*. *X_i_* represents the count of rCNVs (deletions, duplications, or total duplications and deletions within a gene-set). *B*_j_represents the corresponding coefficient for covariate *j* (e.g, age, sex) for individual *i* and *Z_i_*_j_ represents the value for the covariate *j* for individual *i*.

This gene-set burden formula can be broken down for the sake for clarity. To be explicit, the core framework of the gene-set burden calculation remains identical between both equations, differing only by the addition of age and sex covariates. In order to directly compare results to previous work^15^, we utilized Formula 2. However, given the role of age and sex in PTSD and trauma exposure we included these in our main model (Formula 3).

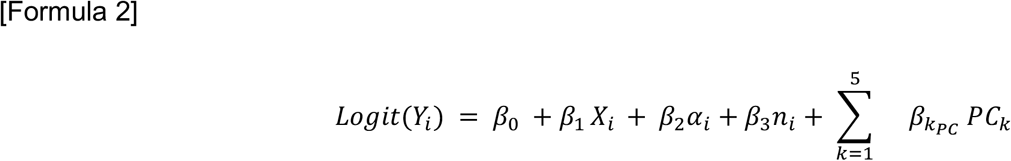

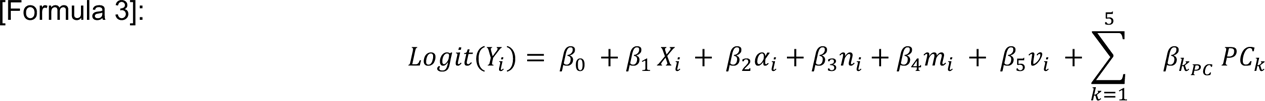

*Y_i_* represents the case/control status. *χ_i_* represents the count of rCNVs (Deletions, Duplications, or Total duplications and deletions within a gene-set) within the target gene set. *β*_O_ represents the intercept. *α_i_* represents genome-wide rCNV count. *n_i_* represents genome-wide average rCNV length (Mb). Genome-wide rCNV count and genome-wide rCNV length (Mb) are added to ensure that the gene-set signal is independent of global burden^52^ (Raychaudhuri et al. 2010). *ν_i_* represents age at DNA collection. *m_i_* corresponds to genetically inferred sex. *PC_k_* corresponds to the first five within ancestry principal components.

#### 3.2.1 Neurodevelopmental (NDD) gene-sets

In an effort to replicate previously published work, we utilized previous NDD gene-sets^15^. NDD CNVs are associated with heterogeneity in clinical presentations (including psychiatric symptoms) suggesting incomplete penetrance^26,64,65^. The NDD gene-sets (N = 46) consisted of 23 gene-sets related to neurofunction or nervous system, 6 brain expression from BrainSpan consortium, a set of loss-of-function intolerant genes as defined by gnomAD v2.0^66^, and 16 brain-expressed gene-sets from human neocortex scRNA data^67^were used in gene-set analysis. We examined whether rCNV burden on NDD gene sets^15^ was associated with PTSD status. Previous rCNV work has commonly relied on rCNVs called from array data, which restricts rCNVs to those with a length of 10 kb and greater. To provide a more accurate comparison to Maihofer et al. (2022) we restricted rCNVs to those with a total length greater than 10 kb. We note that while 46 gene sets were specifically related to NDD, 7 gene sets were labeled as negative control gene sets. However, given the overlap of several of the negative mouse control gene-sets (e.g., cardiovascular [PhMm_Aggr_CardvascMuscle_all], endocrine/exocrine processes [PhMm_Aggr_EndoExocrRepr_all]) associated with stress pathophysiology^68,69^ we selected an additional negative control gene set composed of 397 housekeeping genes to assess rCNV burden^29^ across cases and controls. For simplicity we refer to the total count of NDD gene sets as 53. First, we followed the same gene-set analysis protocol as Maihofer et al. (2022) (Formula 1). Therefore, to account for multiple testing across gene sets (N=204) and three rCNV metrics (deletion count and duplication count), we applied the Benjamini-Hochberg (BH) False Discovery Rate (FDR) procedure^28^. Results were considered significant at an FDR *P* < 0.05.

#### 3.2.2 Common-variant based gene-sets

Cell type specific expression within the dorsolateral prefrontal cortex is associated with PTSD risk^70^. As a result, we also included significant (FDR < 0.05) eQTL SMR genes of single cell types within the dorsolateral prefrontal cortex^7^(Supplementary Table 19). PGC gene-set lists utilized here were generated using European individuals. However, because all five gene sets were based on expression level data, and given the high consistency in direction and magnitude in gene expression^71^ across ancestral groups, we conducted gene-set analyses across AFR and AMR groups as well. In addition, we utilized 146 mouse mutant gene sets previously associated with neuropsychiatric disorders^16–18^. Gene sets were largely implicated in abnormal learning/memory, conditioning, and brain/nervous system development. This method relies on rCNV count assess the frequency and load of rCNV events while also correcting for total genome-wide burden.

A total of five gene-sets derived from the most recent PGC PTSD GWAS^7^ were tested (Nievergelt et al., 2024, Supplementary Table 5 and 6) for rCNV burden across count and length predictors. Protein coding genes^7^ (Nievergelt et al., 2024, Supplementary Table 9), gTEX brain tissue derived TWAS^7^ (Supplementary Table 16), and eQTL SMR^7^ (Nievergelt et al., 2024, Supplementary Table 17) gene sets were selected.

As with the NDD gene-set analyses, multiple testing was controlled using the False Discovery Rate (FDR) method, with results considered significant at an FDR-adjusted *P* < 0.05. The FDR adjustments were applied separately within each testing framework based on the number of predictors multiplied by the number of evaluated gene-sets (N=204).

### 2.0 Supplemental Results

#### 2.1 CNV Characterization

We conducted all analyses using a set of rCNVs previously called by AoU. Importantly, all CNVs were filtered for quality on an individual basis using whole genome dosage. Samples whose coverage across the genome was highly variable were subsequently removed by AoU. Using this set of CNVs passing AoU quality control measures^2^ we selected those CNVs with a population specific MAF < 1% across two thresholds 1) no length thresholds and 2) 10 kb length threshold.

In order to provide a complete baseline, we examined rCNV frequency across a global MAF of < 1%. A total of 942,179 rCNVs across all lengths were selected including 693,060 deletions and 249,119 duplications were observed with a MAF cutoff of 1%. Deletion length (median: 0.76 kb) was not significantly larger than duplication length (median: 0.29 kb) (Mann-Whitney *U* = 91491743844.0, *P* = 1.0). In an effort to replicate prior CNV work by the PGC PTSD^15^ we then restricted rCNVs to those greater than 10 kb to carry out all genome-wide and geneset burden analyses. A total of 124,271 rare duplications and deletions (> 10 kb) were selected, including 71,340 deletions and 52,931 duplications. The median CNV size was 28.34 kb, with the median deletion size (23.13 kb) being significantly smaller than the median duplication size (37.74kb) (Mann-Whitney *U* = 14.68 x10^8^, p-value was below the point of numerical precision)

As a sensitivity analysis we characterized the between population (EUR-like, AFR-like, AMR-like) rCNV frequency of PTSD cases and controls using a Fishers Exact Test within PLINK 1.9 (Supplementary Table 9). We identified a significant divergence in rCNV frequencies between AFR-like and EUR-like cohorts (*P* < 0.001), with the AFR-like cohort exhibiting a 2.6-fold higher mean frequency across ∼50,000 variants. Similarly, the AMR-like mean of rCNVs was 1.98x higher than AFR-like. This justifies our ancestry specific modeling and meta-analysis in order to minimize false positives. Given this, we filtered rCNVs based on ancestry specific MAF for all analyses presented.

#### 2.2 rCNV Region Level Analyses

##### 2.2.1 rCNVs (MAF < 1% and > 10 kb)

Filtering for rCNV’s with an ancestry specific allele frequency less than 1% and greater than 10 kb yielded no significant results in any ancestry group. Two nominal associations were found in EUR in the cytoband 2p22.3. The mean rCNV length across these nominal rCNVs approaching was 19,818 bp and genomic inflation was controlled (lambda_1000_ = 0.93).

Given the MAF and sample size, genomic control was deflated in AFR (lambda_1000_ = 0.76). This indicates we were underpowered to detect significant rCNV associations. Four nominal rCNV associations in AFR were found with 16p23.1 as the most significant rCNV (*P* = 3.91 x 10^-5^, *P_BF_* = 0.11, OR = 7.54). This 39,909 bp region overlapped several genomic features, including *TMEM231P1*, *CHST5*, ENSG00000291051, and ENSG00000260092.

Six (4 deletions, 2 duplications) rCNVs were nominally significant in AMR, however genomic control was significantly deflated in AMR (lambda_1000_ = 0.844). The most significant rCNV included duplication 10q11.22 (*P* = 4.74 x 10^-5^, *P_BF_* = 0.14, OR = 4.78) overlapping the long intergenic non-protein coding RNA, *AGAP4,* with a length of 10735 bp.

#### 2.3 rCNV Gene-Set Burden Analyses

##### 2.3.1 PTSD Common Variant Gene-Set Results

The regression results (P-values and effect sizes) for the ‘All Lengths’ and ‘10kb+’ analyses utilizing PTSD PGC common variant gene sets were exactly identical within the EUR, AFR, and AMR cohorts (Supplementary Table 5 and 9). This identity occurs because the extra, small variants (<10kb) did not identify any new individuals as carriers. Instead, they overlapped the targeted gene sets in the exact same individuals who already carried a larger macro-structural variant (>10kb) at those genomic locations.

PTSD common variant prioritized gene-set burden results yielded identical association statistics across all rCNV length and size-restricted (≥10kb) tiers (Supplementary Table 5 and 9). While neither configuration achieved strict statistical significance under our current sample size, this absolute symmetry indicates that the localized rare variant architecture within this specific compact gene set is driven entirely by macrostructural carriers, with no sub-10kb variants overlapping these target loci. The PTSD common-variant prioritized gene sets were significantly restricted in gene count (Supplementary Files) leading to a massive reduction in mutational target size compared to broader tissue-specific pathways. For example, while the single-cell dorsolateral prefrontal cortex set (brain_tissue_sc_dlPFC_suppl_tbl_19; ∼615 genes) captured 12,718 rare variants in the AFR-like cohort and 18,425 in the EUR cohort, the core post-GWAS prioritization set (magma_suppl_tbl_13; ∼179 genes) captured only 31 and 46 rCNVs, respectively. When restricting analyses to rCNVs greater than 10kb, these counts collapsed to just 12 variants in AFR and 21 in EUR. This extreme scarcity of structural overlapping carriers demonstrates that the lack of significance within these targeted pathways.

### 3.0 Supplementary Information

#### 3.1 Extended Analysis of Rare CNV Discrepancies and Implications

rCNVs (MAF < 1%) have been implicated across several psychiatric and neurodevelopmental disorders^15,26^. Although CNVs have been less studied with regard to PTSD, one prior CNV study^15^ identified significant neurodevelopmental gene-sets associated with PTSD status with array-based data. Prior work investigating rare structural variation has often been limited by the use of array based CNV calling and the sole reliance on EUR-like inferred ancestry samples. Therefore, the current study analyzed rare copy number variation from sequencing data among all available ancestry samples using a large-scale population representative biobank dataset. Given the absence of prior work utilizing sequencing based rCNVs or rCNVs across additional ancestries, this limits our ability to directly contextualize the results against previous studies.

#### 3.2 Comparison to PGC and implications of rCNVs from sequencing data

Two additional factors may contribute to discrepancies in results from the prior PGC work^15^ including sample size and phenotyping definition. The current analyses relied on a case sample size substantially smaller than the case sample size in this work. In addition, we relied on a narrow EHR definition of PTSD in order to capture PTSD cases in a population biobank. Prior work has often used an inclusive definition that grouped PTSD with Acute Stress Disorder, Adjustment Disorders, and other non-specific stress reactions, potentially increasing phenotypic noise that may introduce non-specific genetic associations. However, we hypothesized that a more strictly defined case definition would be more likely to be enriched for rare variation. This hypothesis is consistent with common variant biobank research focusing on broadly defined MDD which displayed non-specific genetic associations, while restricting MDD case status to a more stringent threshold increased the specificity of genetic signal to MDD^20^. Consequently, minimal phenotyping may bias views on the genetic architecture of PTSD, along with the relevance of rare CNVs and biological pathways.

#### 3.3 Further Discussion of Limitations

Significantly the current results utilized binary case/control status derived from EHR (within meta-analysis and within ancestry), in comparison to prior work which primarily utilized continuous or ordinal symptoms severity scales^15^. This suggests that EHR derived cohorts with an explicit diagnosis of PTSD may represent a more penetrant clinical phenotype. This potential enrichment for severe cases may counter the loss of statistical power from relying on a binary, rather than continuous phenotypic measure.

The current results here should be considered in the context of several limitations. Defining case and control status using EHR data is prone to bias. Specifically, we were unable to be certain controls are true controls, as not all patients are screened for PTSD. For example, we observed between 2.4-3.5% of controls having at least one acute stress disorder and it is possible a portion of these individuals transitioned to a PTSD diagnosis. Other biases include an uneven distribution of healthcare utilization and avoidance itself being a criterion of PTSD, therefore it is possible we are missing cases. Given the abundance of comorbidity and patterns in provider-given diagnostic codes, sample overlap with other internalizing conditions is unavoidable. Consistent with known clinical heterogeneity of PTSD^72^, psychiatric comorbidity was highly prevalent in the cases. A large proportion of PTSD cases also carried diagnoses of anxiety disorders and major depressive disorder. Finally, EHR adjustment disorder cases were not included here, as done in some prior work examining a broader PTSD definition^7^. Although adjustment disorders are considered on the stress continuum and closely related to PTSD^73^, our analysis focused exclusively on cases meeting a narrow definition of PTSD, as we hypothesized that this more stringent phenotype represents greater symptom severity and, consequently, a higher likelihood of rare variant enrichment.

Additionally, SVs remain difficult to classify as they tend to reside within repetitive DNA, which makes their characterization more difficult^50^. Along with this, coverage may not be uniform across the genome which is a limiting factor in paired read SV calling methods^74^. For example, longer genes will have better average coverage compared to smaller genes. However, SV calling methods such as those accounting for read depth (e.g., Event Wise Testing) can account for these biases^75^. Despite the use of GATK-SV, an ensemble method utilizing read depth approaches, validation remains challenging. This is particularly relevant, given that intergenic rCNVs comprise approximately 40% of our dataset.

---

**Supplementary Figure 1.**
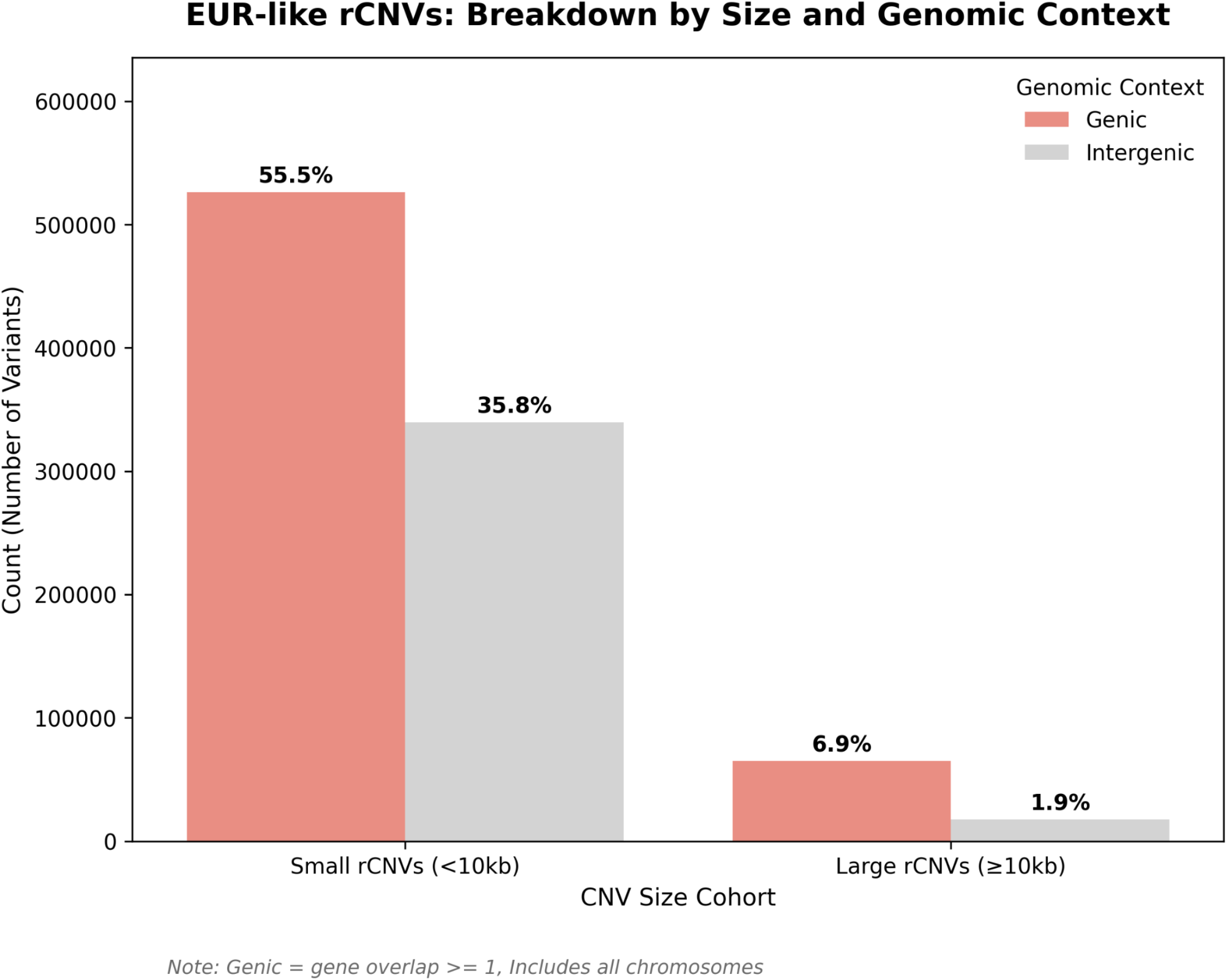
EUR-like rCNVs Breakdown of rCNVs by Size and Genomic Context. Proportion of EUR-like rCNVs intersecting genic versus intergenic regions across small (<10kb) and large rCNVs (>=10kb). A minimum overlap of 1bp was required to classify a rCNV as intersecting a genic region.

**Supplementary Figure 2.**
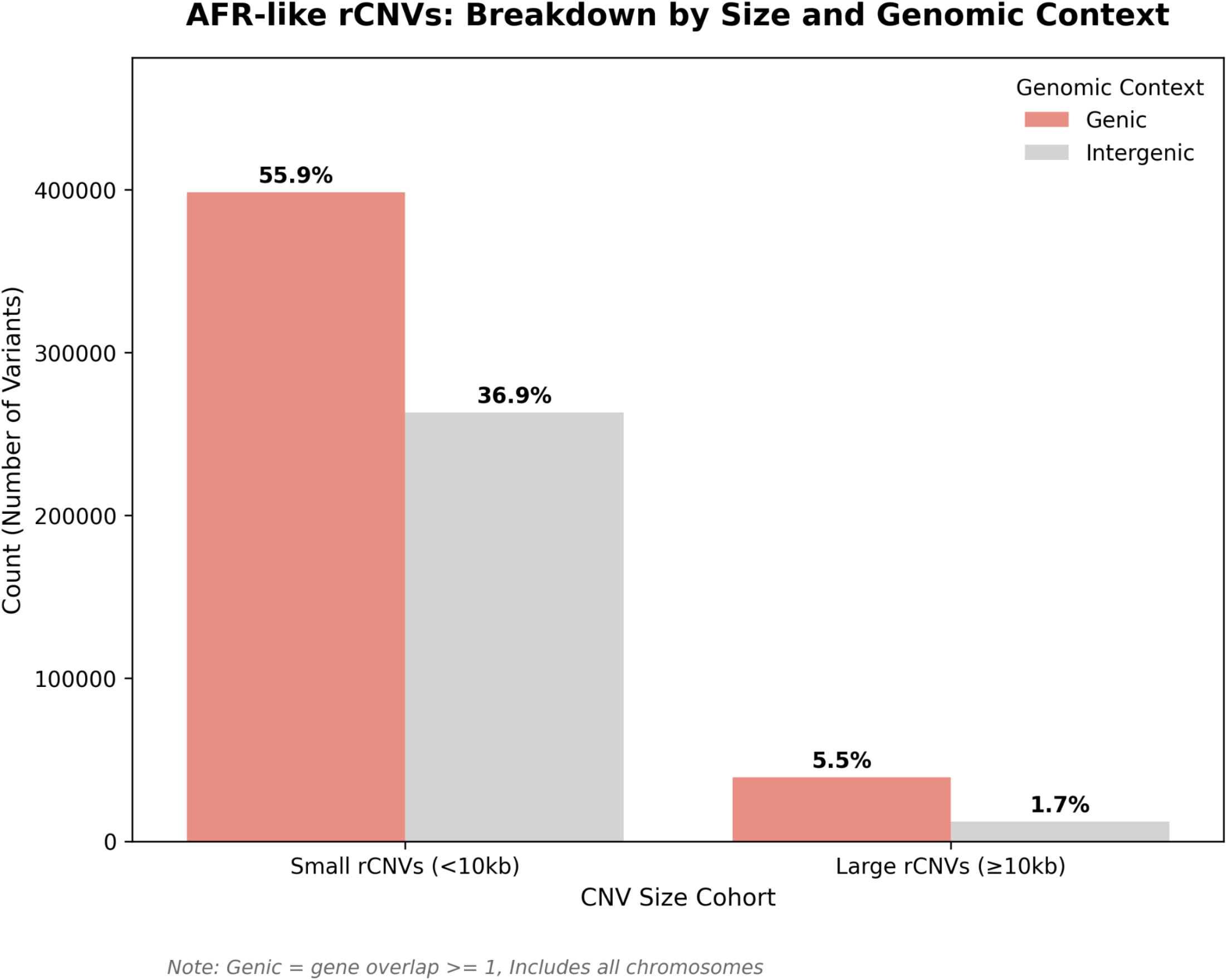
AFR-like rCNVs Breakdown of rCNVs by Size and Genomic Context. Proportion of AFR-like rCNVs intersecting genic versus intergenic regions across small (<10kb) and large rCNVs (>=10kb). A minimum overlap of 1bp was required to classify a rCNV as intersecting a genic region.

**Supplementary Figure 3.**
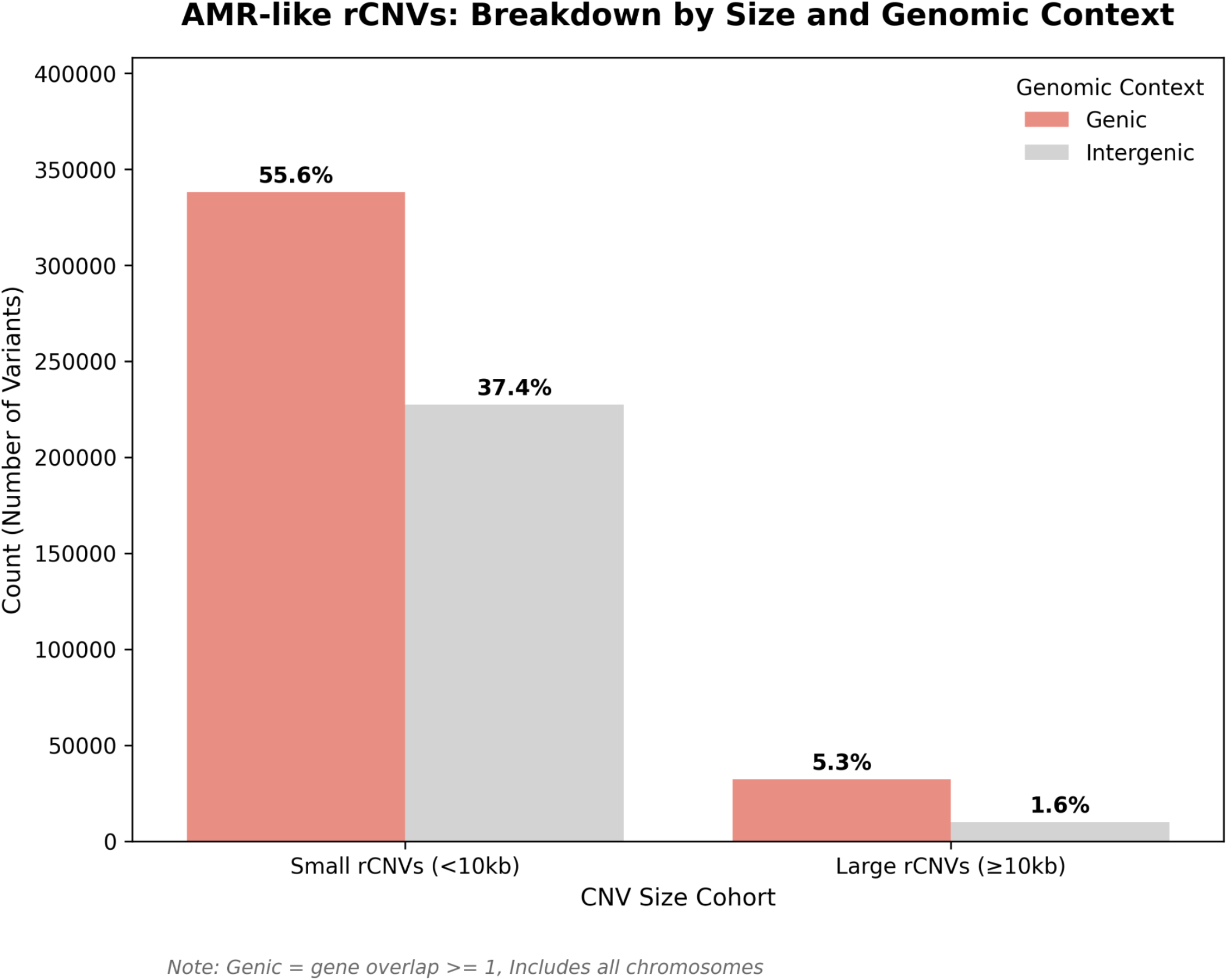
AMR-like rCNVs Breakdown of rCNVs by Size and Genomic Context. Proportion of AMR-like rCNVs intersecting genic versus intergenic regions across small (<10kb) and large rCNVs (>=10kb). A minimum overlap of 1bp was required to classify a rCNV as intersecting a genic region.

**Supplementary Figure 4.**
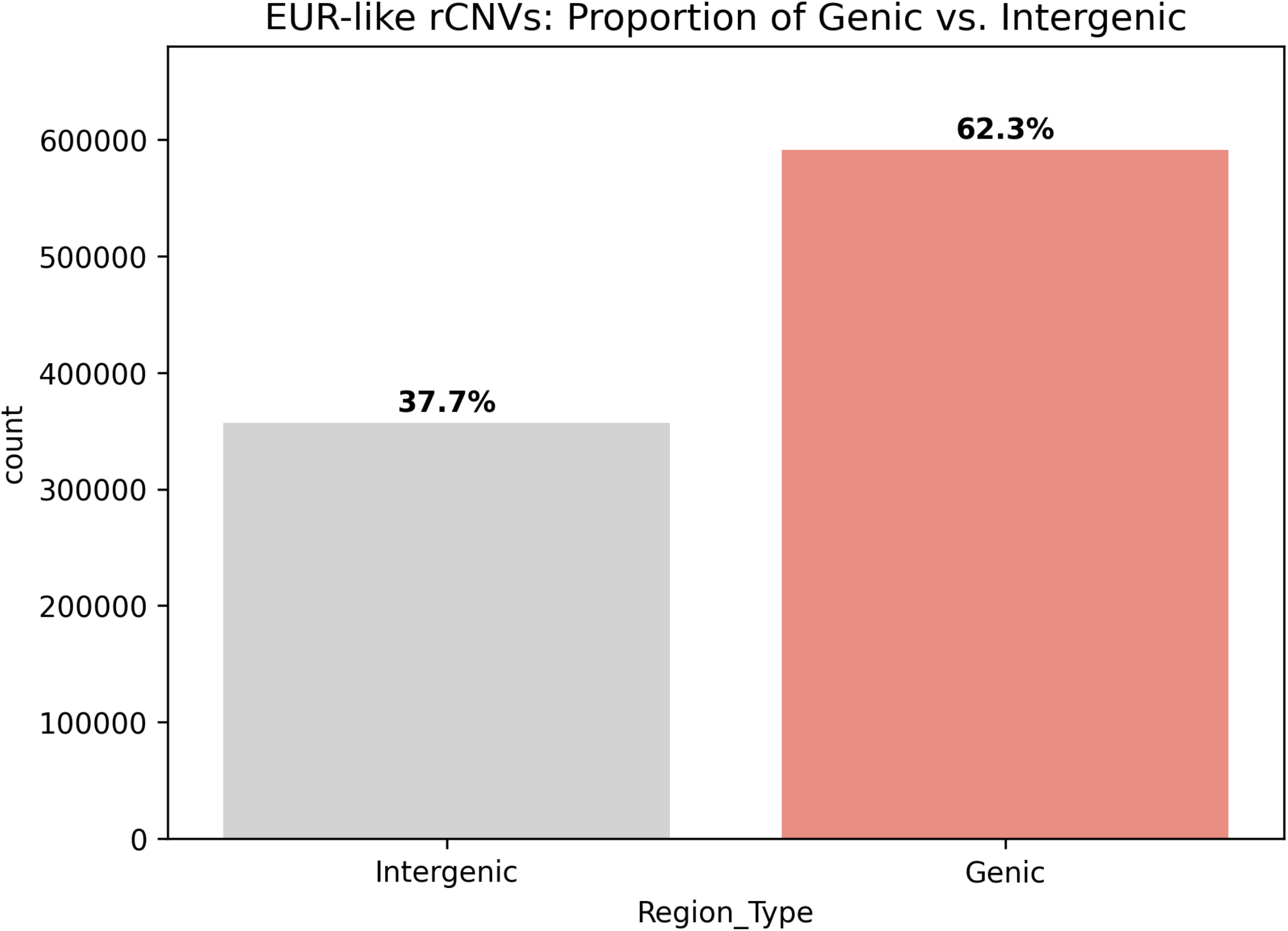
EUR-like rCNVs Proportion of Genic vs. Intergenic. Proportion of EUR-like rCNVs intersecting genic versus intergenic regions. A minimum overlap of 1bp was required to classify a rCNV as intersecting a genic region.

**Supplementary Figure 5.**
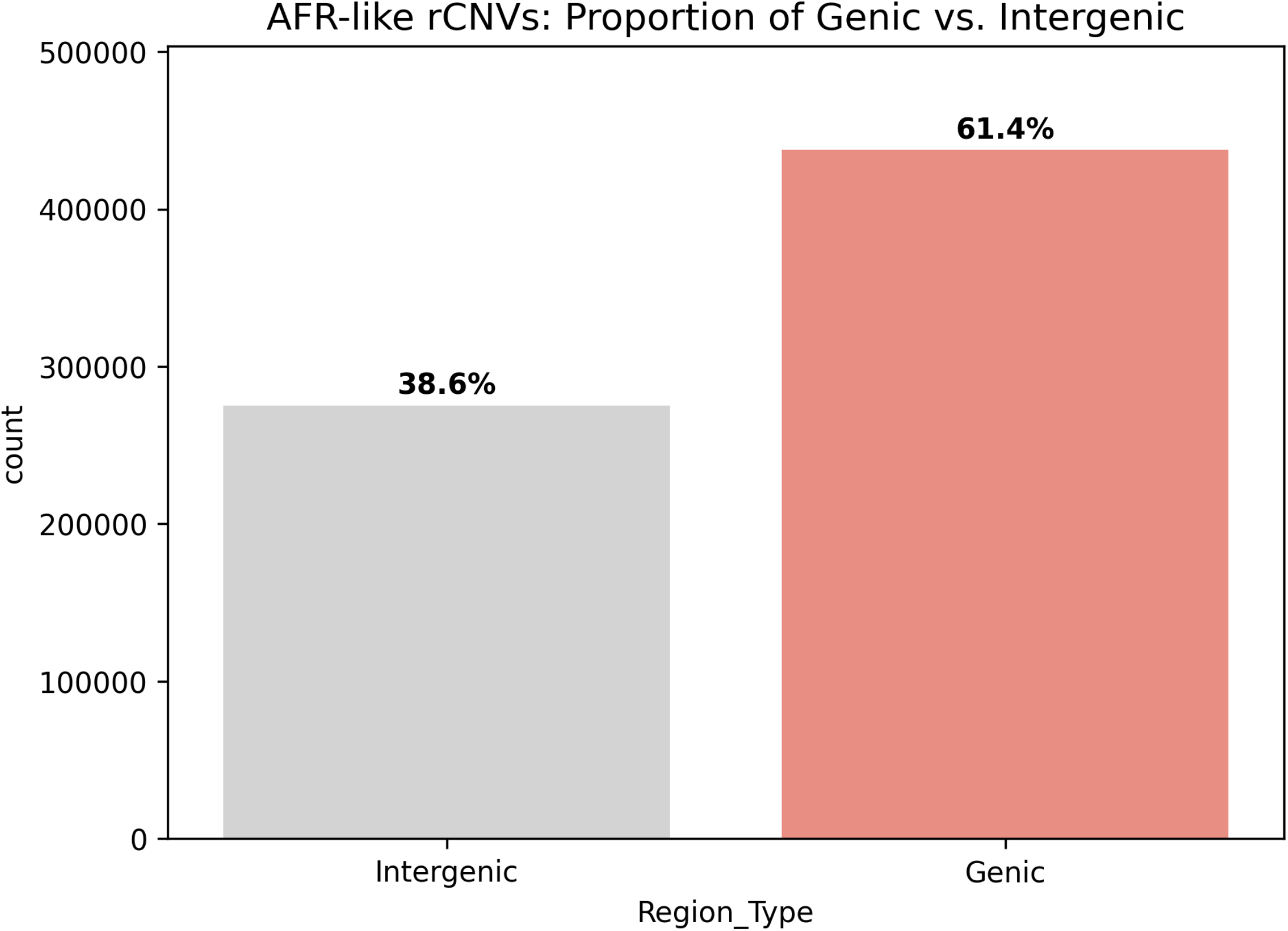
AFR-like rCNVs Proportion of Genic vs. Intergenic. Proportion of AFR-like rCNVs intersecting genic versus intergenic regions. A minimum overlap of 1bp was required to classify a rCNV as intersecting a genic region.

**Supplementary Figure 6.**
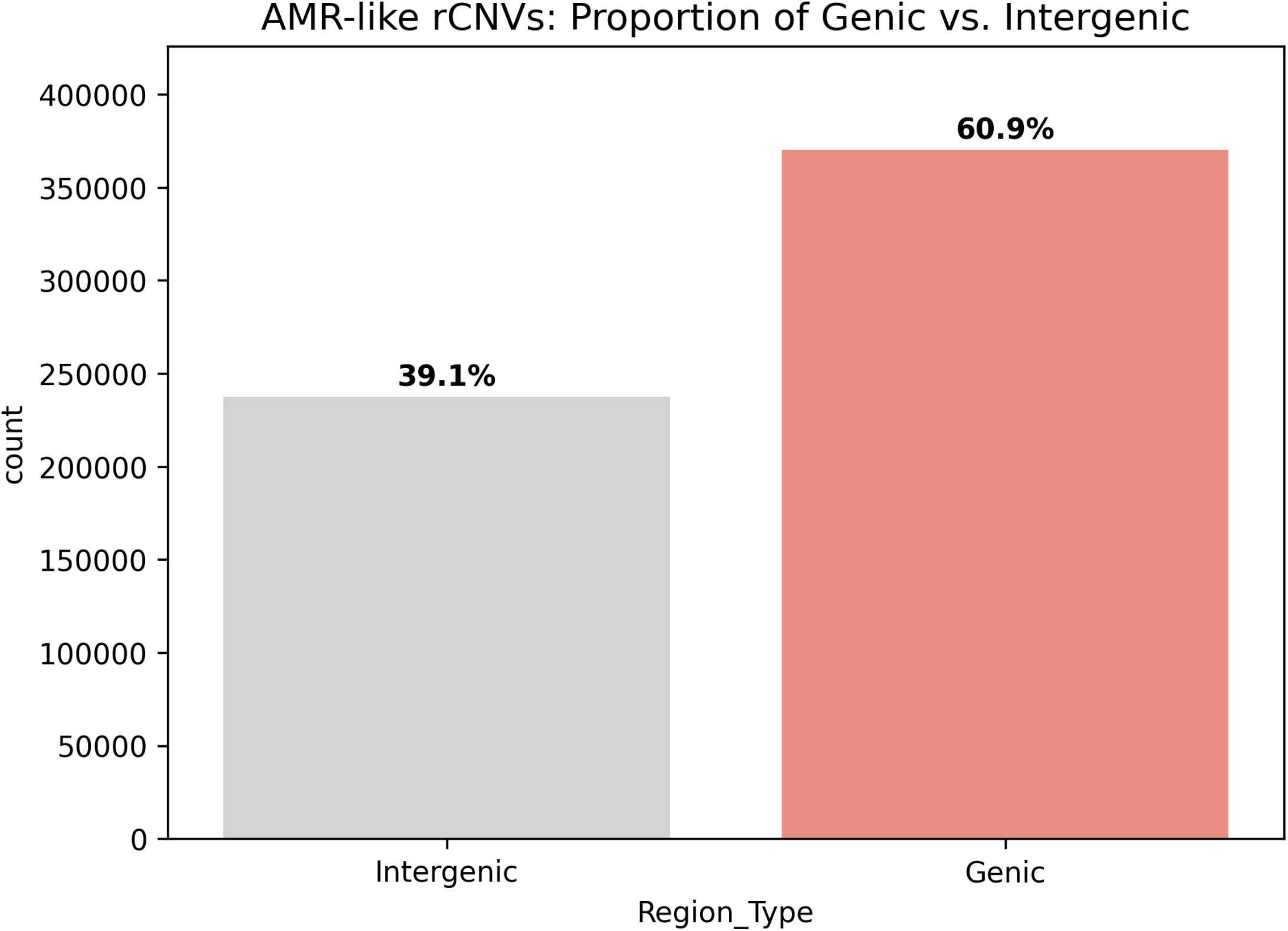
AMR-like rCNVs Proportion of Genic vs. Intergenic. Proportion of AMR-like rCNVs intersecting genic versus intergenic regions. A minimum overlap of 1bp was required to classify a rCNV as intersecting a genic region.

**Supplementary Figure 7.**
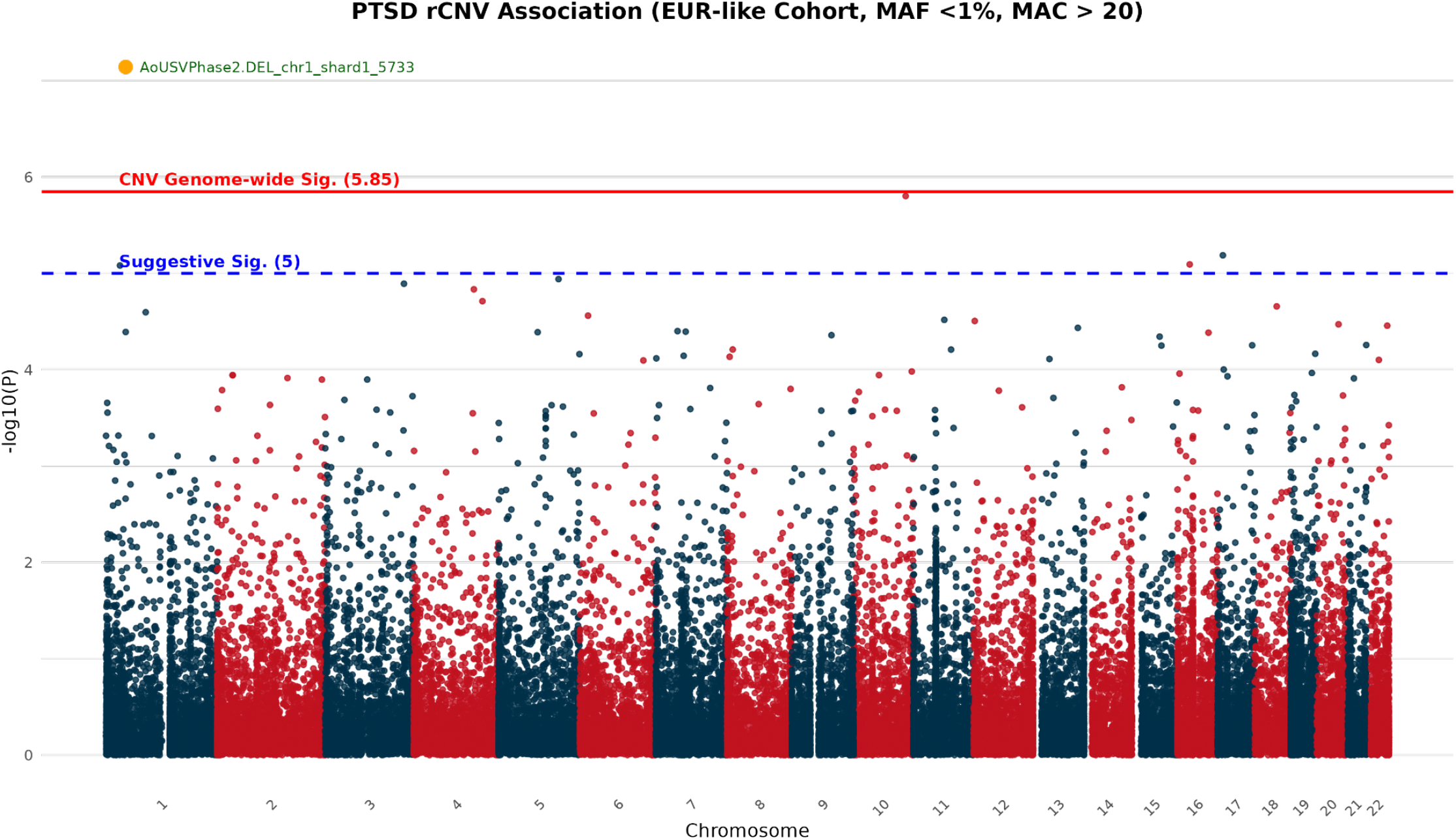
Manhattan Plot rCNV EUR-like. Manhattan plot for Firth’s regression in EUR-like rCNVs (MAF < 1%, N = 35,250) across all rCNV lengths.

**Supplementary Figure 8.**
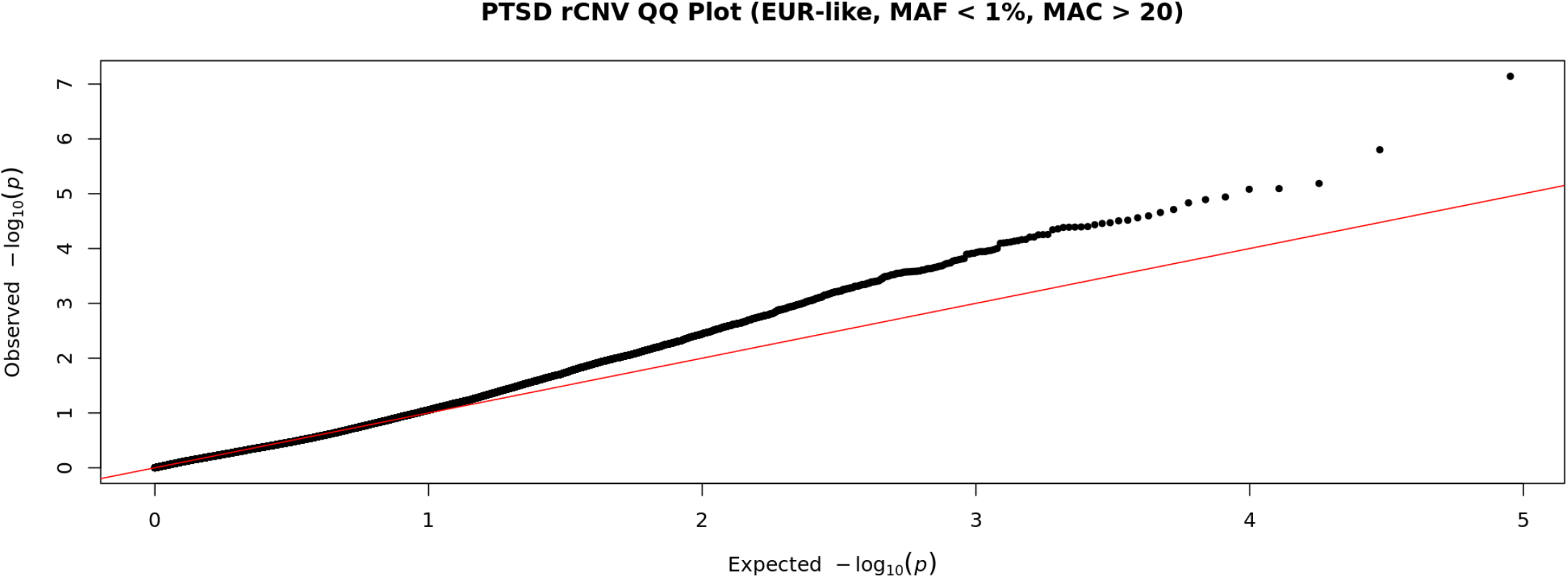
QQ Plot rCNV EUR-like. QQ plot of genomic inflation in Firth’s regression in EUR-like rCNVs (MAF < 1%, N = 35, 250) across all rCNV lengths. Lamaba = 0.95 and Lamba_1000_ = 0.96.

**Supplementary Figure 9.**
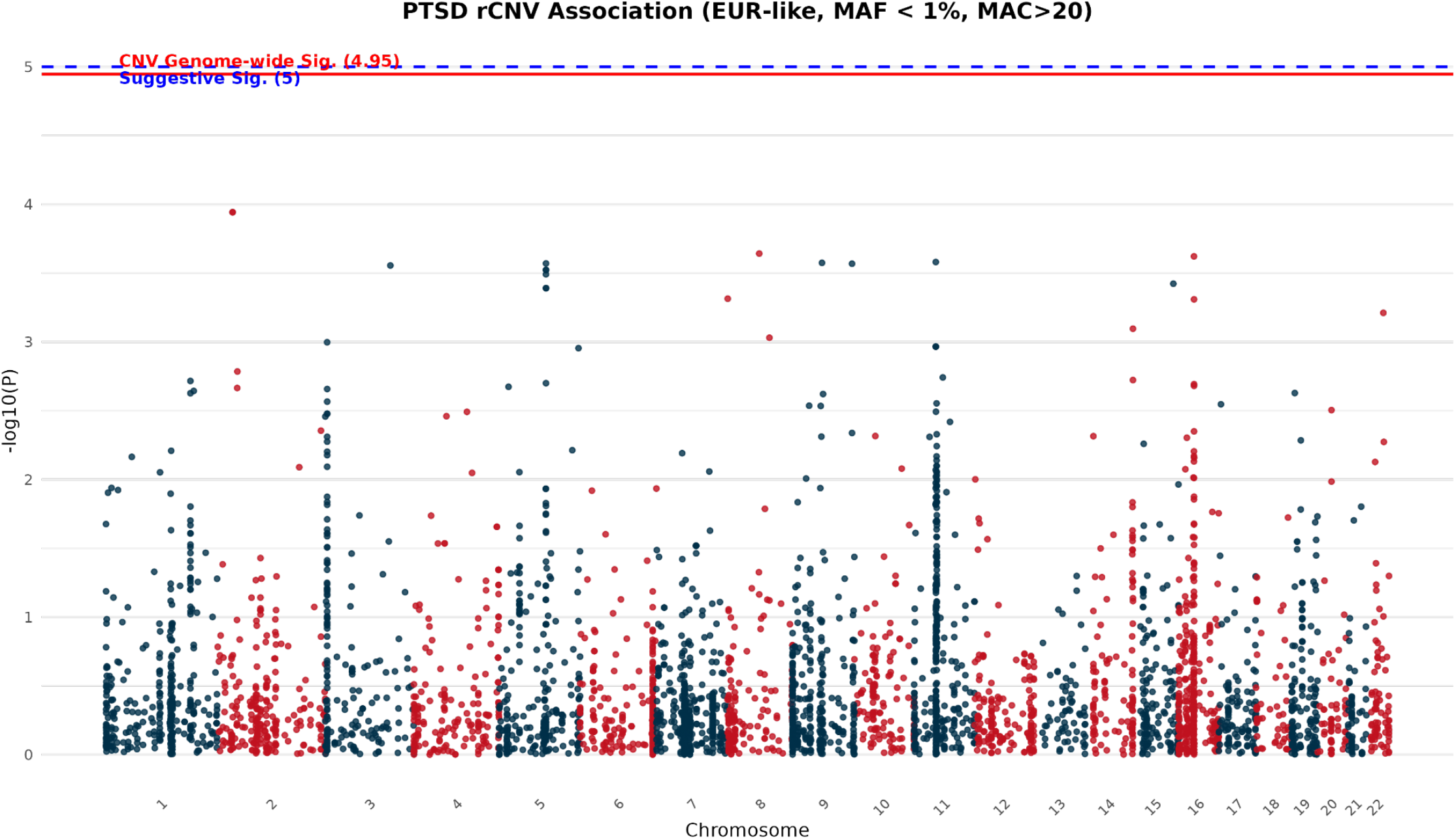
Manhattan Plot rCNV (10 kb) EUR-like. Manhattan plot for Firth’s regression in EUR-like rCNVs (MAF < 1%, N = 4423) across rCNVs greater than or equal to 10 kb.

**Supplementary Figure 10.**
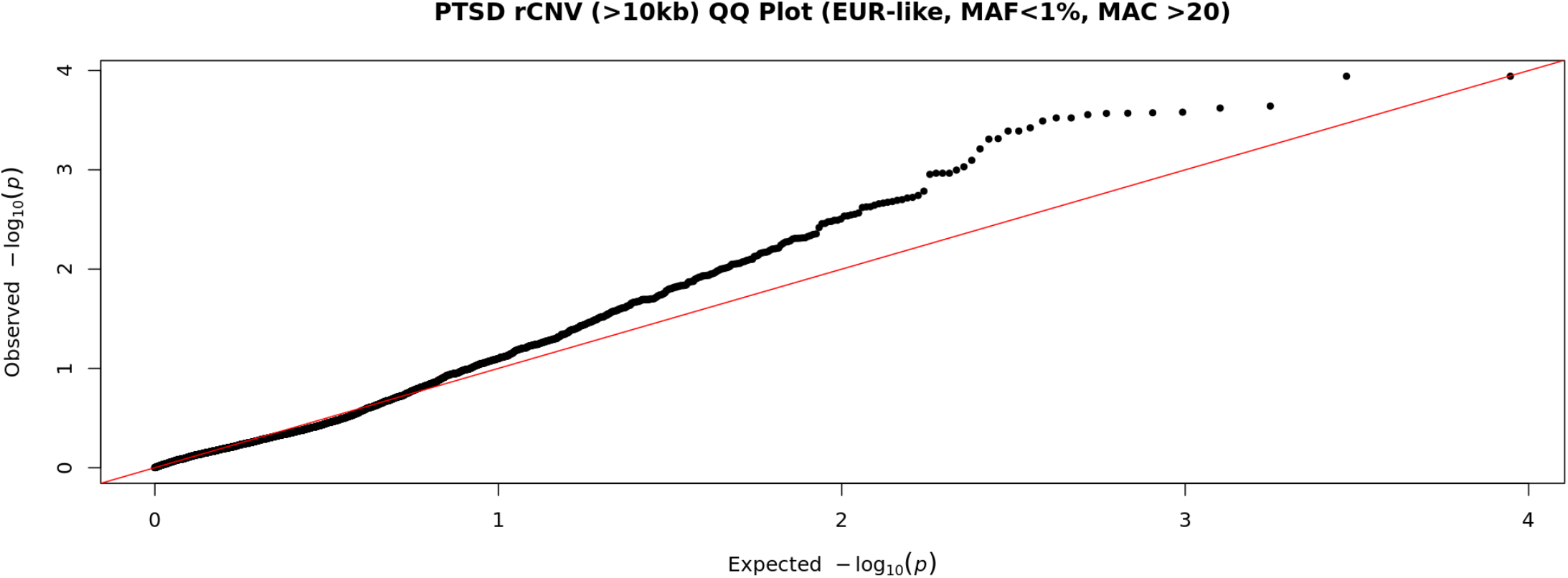
QQ Plot rCNV (>10 kb) EUR-like. QQ plot of genomic inflation in Firth’s regression in EUR-like rCNVs (MAF < 1%, N = 4,423) across all rCNV lengths. Lamaba=0.85 and Lamba_1000_ = 0.93.

**Supplementary Figure 11.**
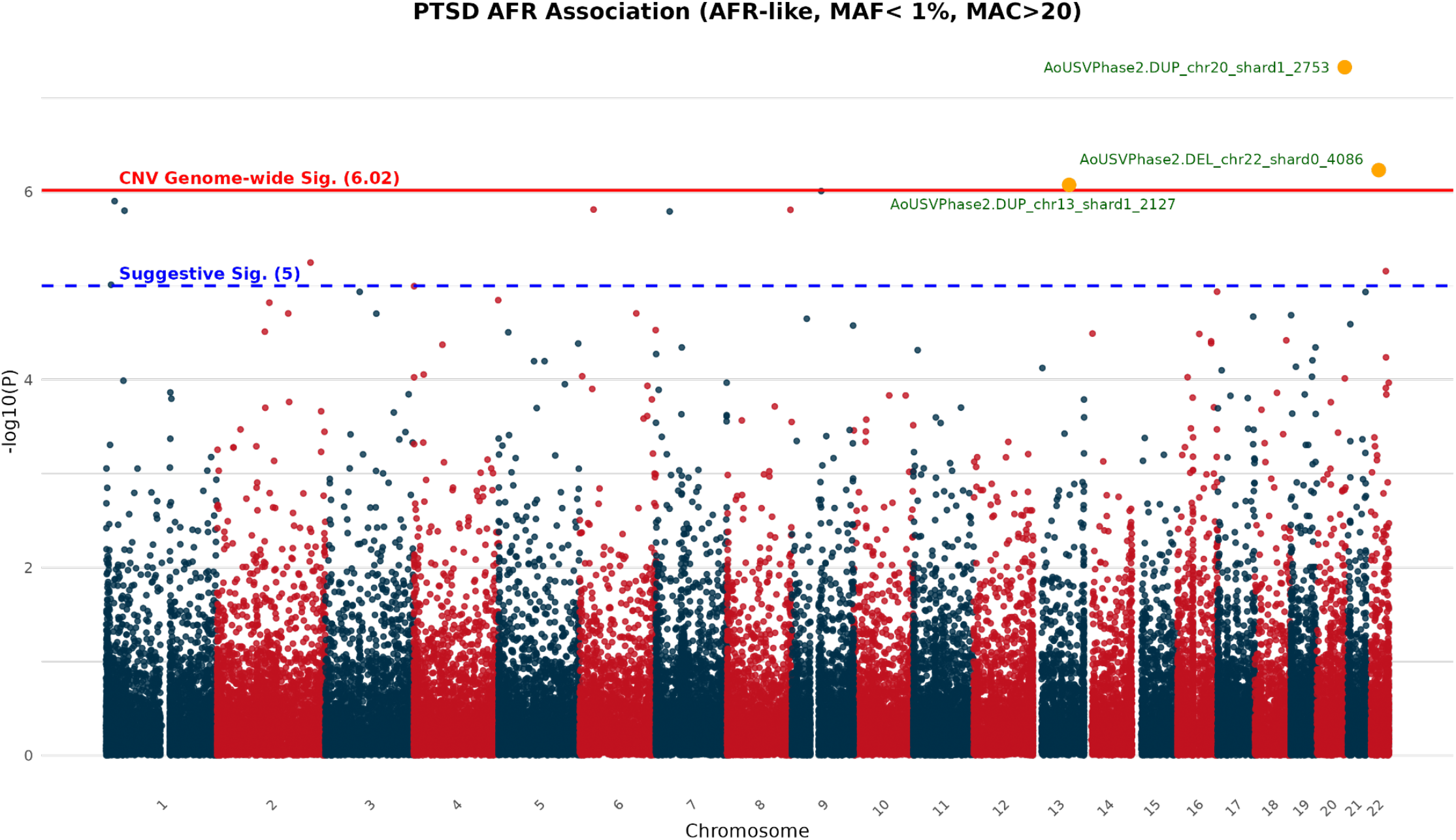
Manhattan Plot rCNV AFR-like. Manhattan plot for Firth’s regression in AFR-like rCNVs (MAF < 1%, N = 51,954) across rCNVs across all lengths.

**Supplementary Figure 12.**
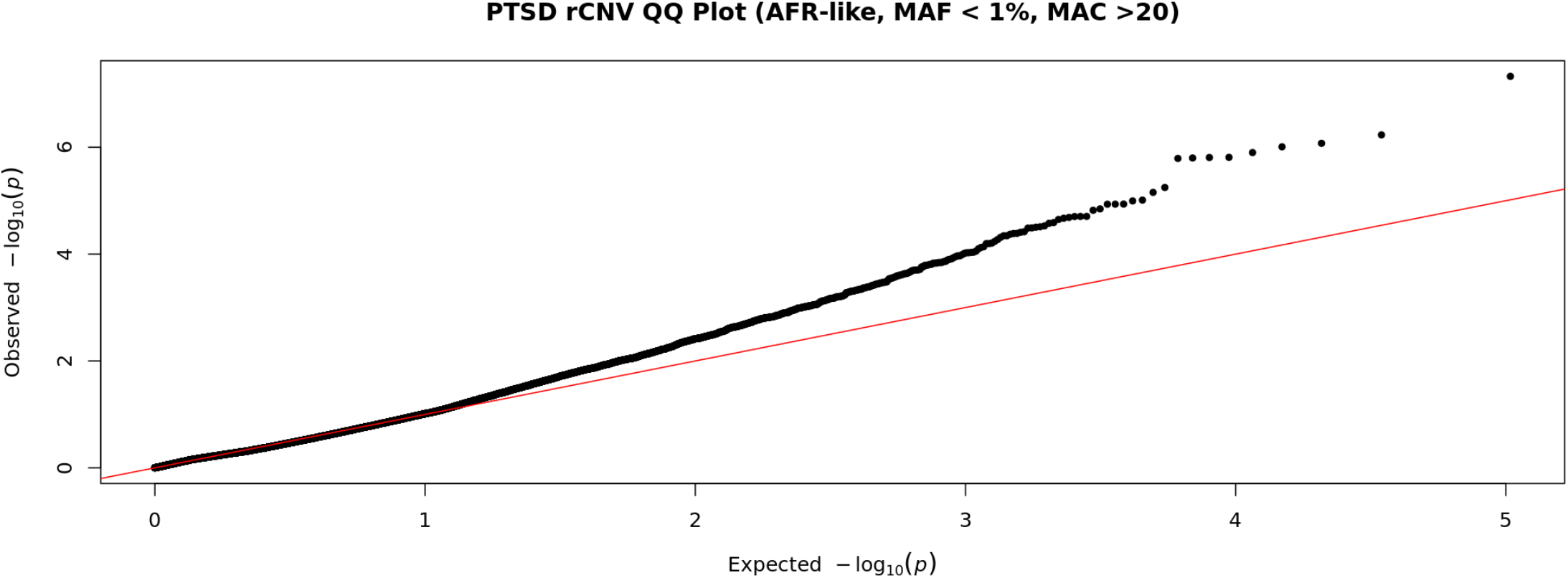
QQ Plot AFR-like. QQ plot of genomic inflation in Firth’s regression in AFR-like rCNVs (MAF < 1%, N = 51954) across all rCNV lengths. Lambda = 0.90 and Lamba_1000_ = 0.90.

**Supplementary Figure 13.**
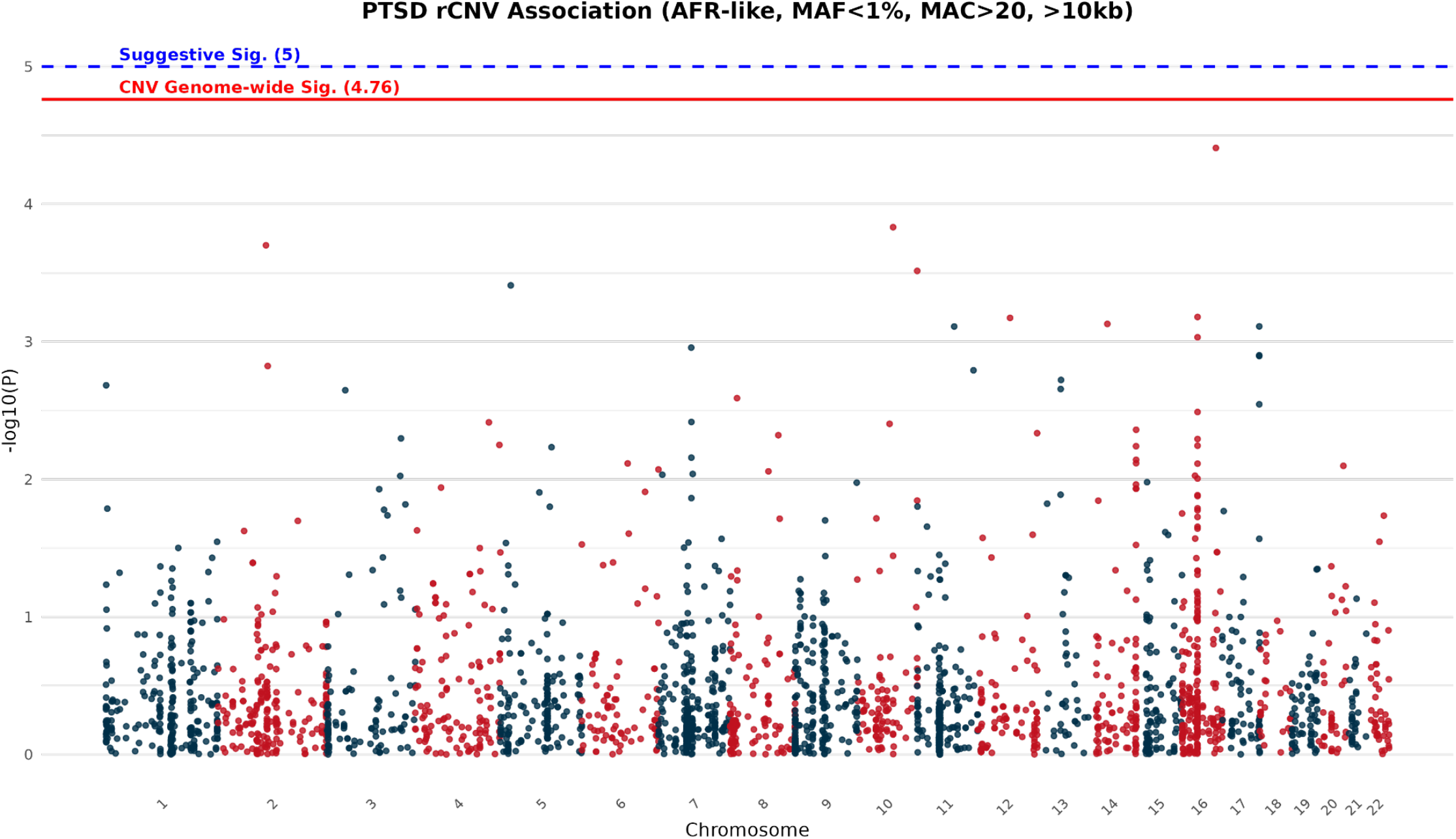
Manhattan Plot rCNV (10 kb) AFR-like. Manhattan plot for Firth’s regression in AFR-like rCNVs (MAF < 1%, 10 kb, N = 2891) across rCNVs greater than or equal to 10 kb.

**Supplementary Figure 14.**
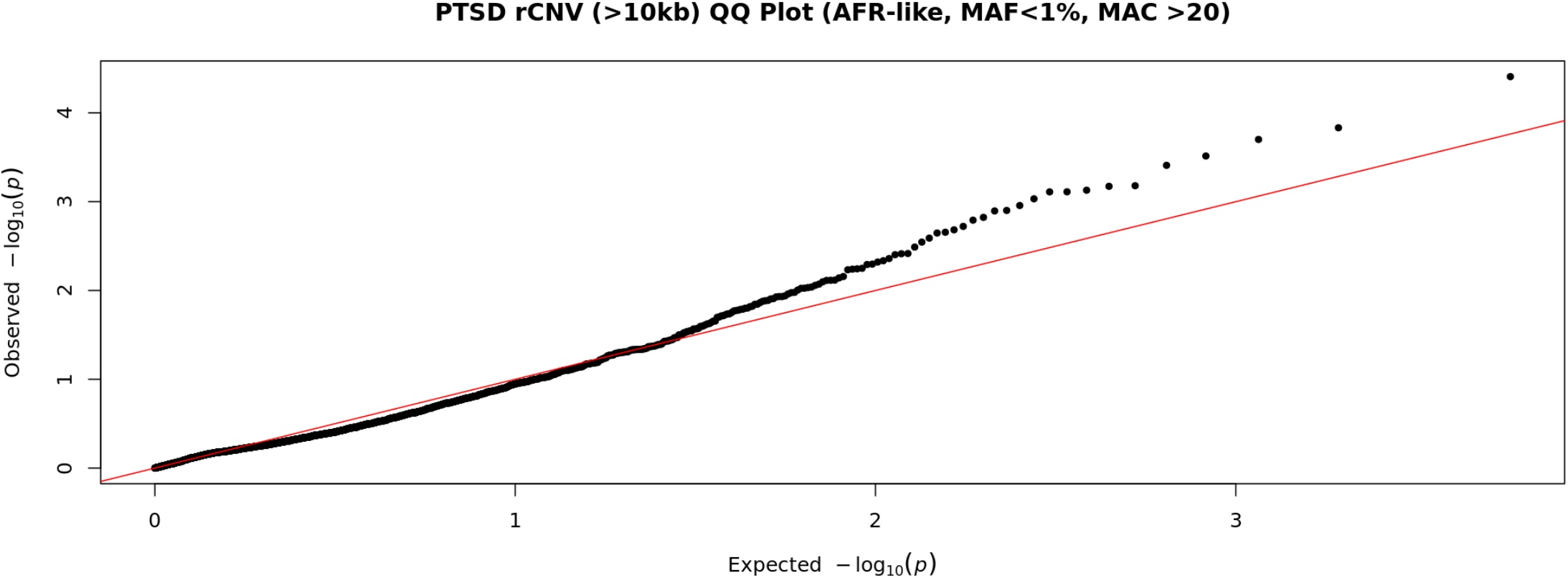
QQ Plot AFR-like (10 kb) QQ plot of genomic inflation in Firth’s regression in AFR-like rCNVs (MAF < 1%, N = 2891) across all rCNVs greater than or equal to 10 kb. Lambda = 0.77 and Lamba_1000_ = 0.76.

**Supplementary Figure 15.**
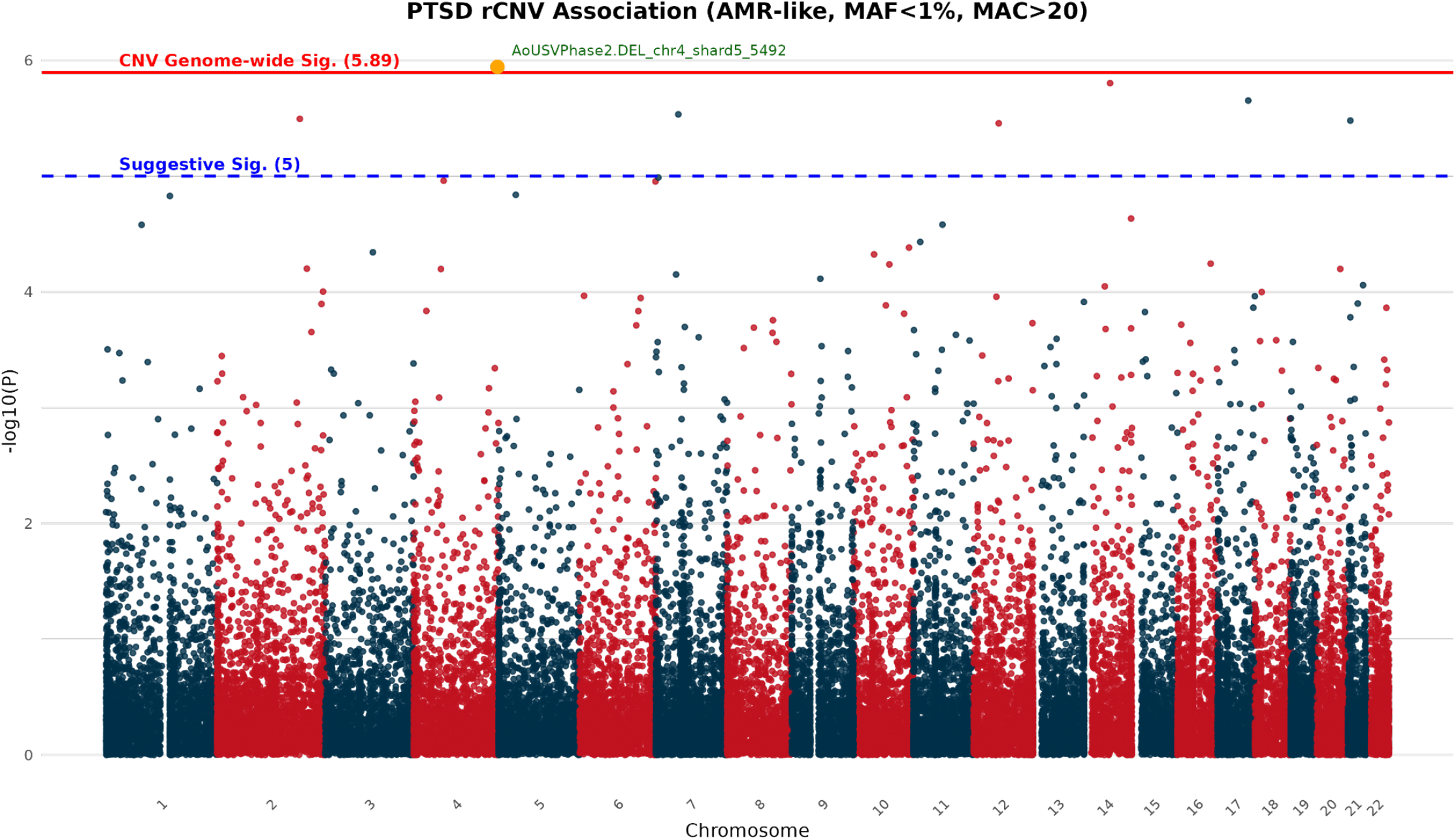
Manhattan Plot rCNV AMR-like. Manhattan plot for Firth’s regression in AMR-like rCNVs (MAF < 1%, N = 39,216) across rCNVs across all lengths.

**Supplementary Figure 16.**
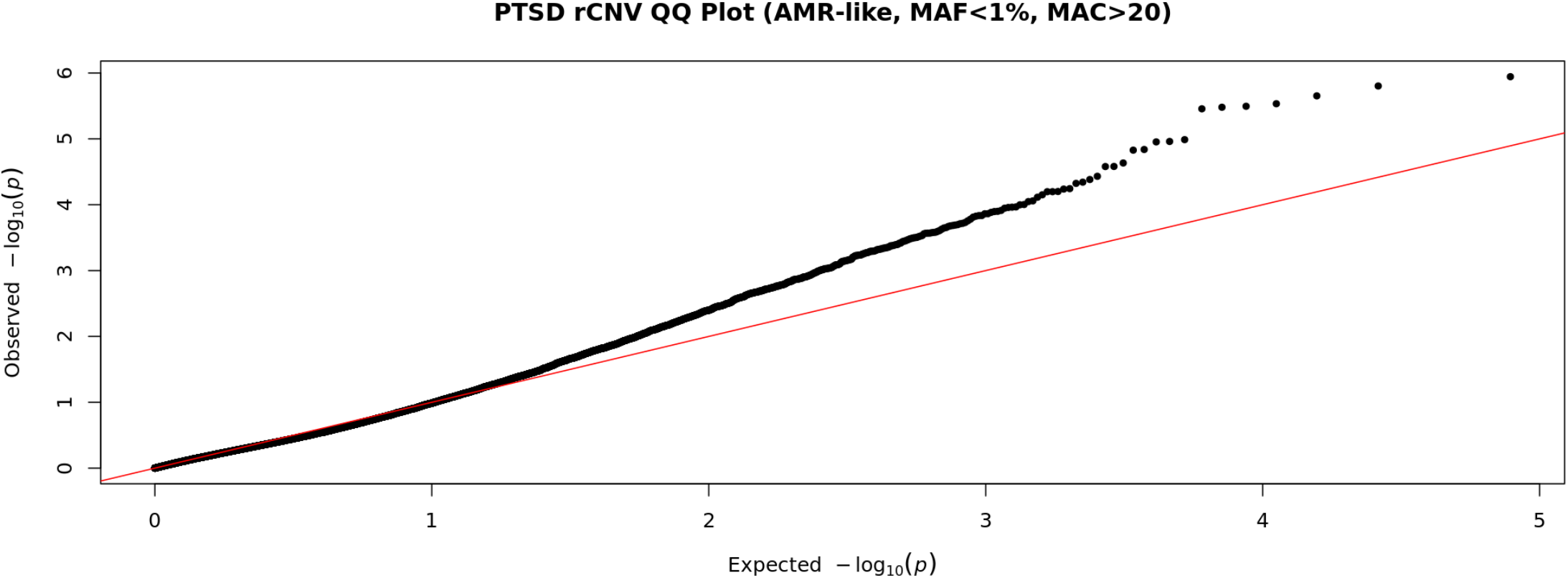
QQ Plot AMR-like. QQ plot of genomic inflation in Firth’s regression in AMR-like rCNVs (MAF < 1%, N = 39, 216) across all rCNV lengths. Lambda= 0.88 and Lamba_1000_ = 0.86.

**Supplementary Figure 17.**
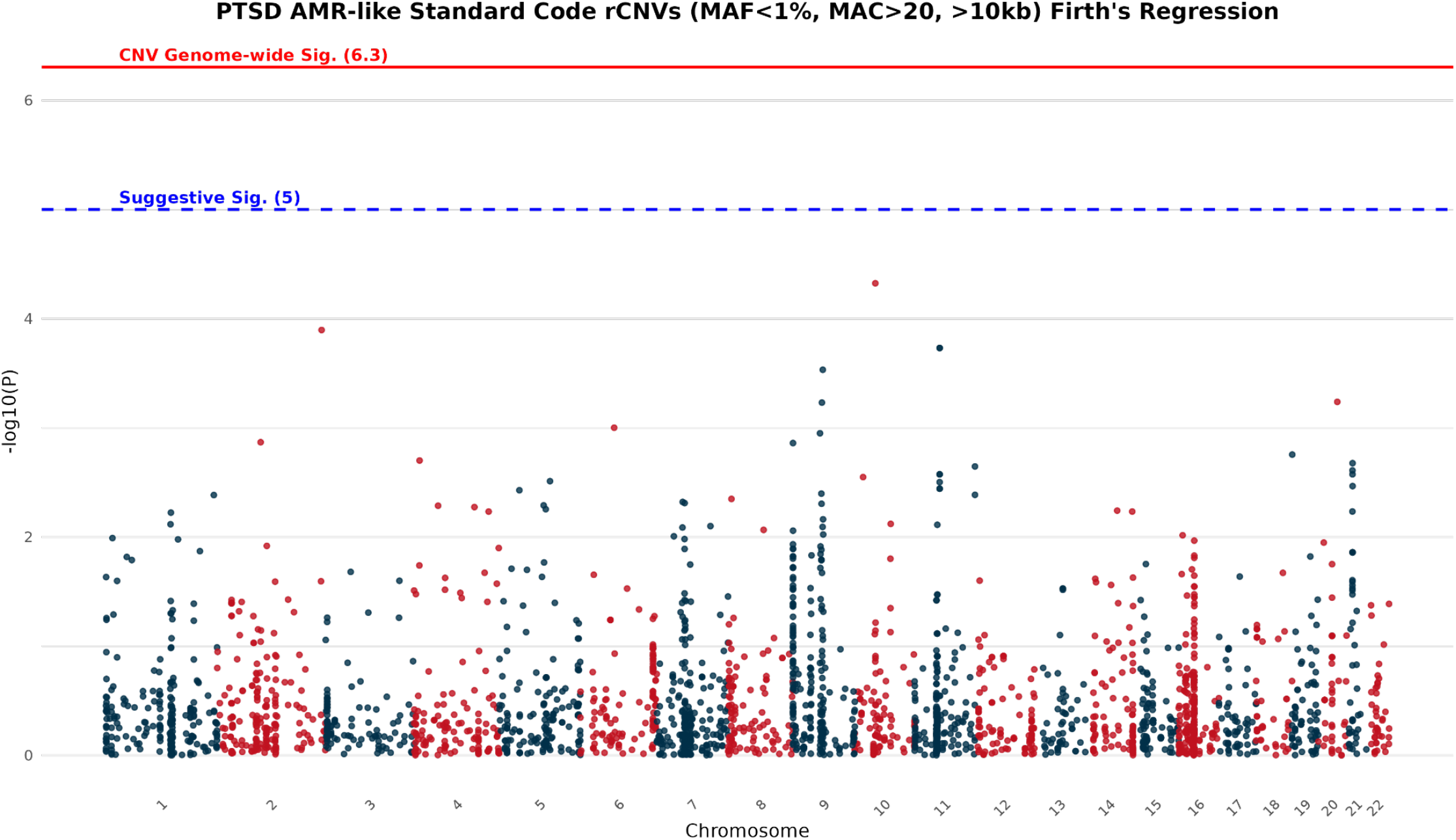
Manhattan Plot rCNV AMR-like (10 kb) Manhattan plot for Firth’s regression in AMR-like rCNVs (MAF < 1%, 10 kb, N=2908) across rCNVs greater than or equal to 10 kb.

**Supplementary Figure 18.**
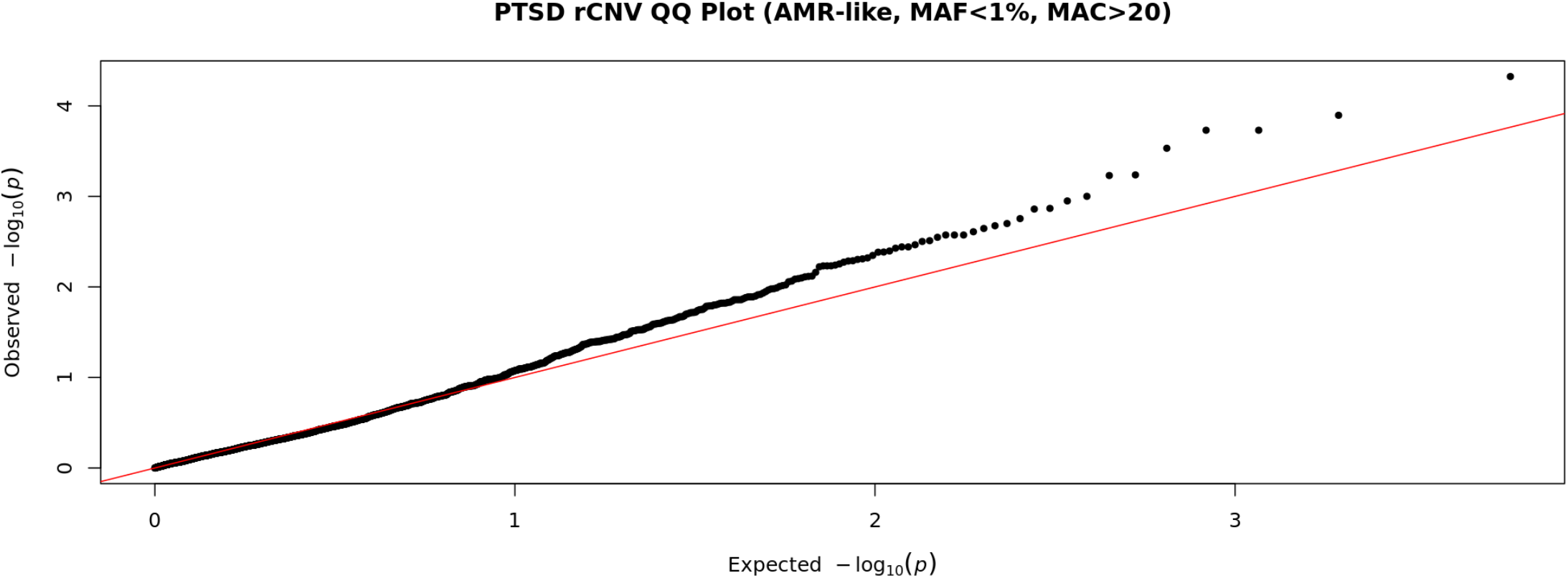
QQ Plot AMR-like (10 kb) QQ plot of genomic inflation in Firth’s regression in AFR-like rCNVs (MAF < 1%, N = 2908) across all rCNVs greater than or equal to 10 kb. Lambda = 0.87 and Lamba_1000_ = 0.84.

**Supplementary Figure 19.**
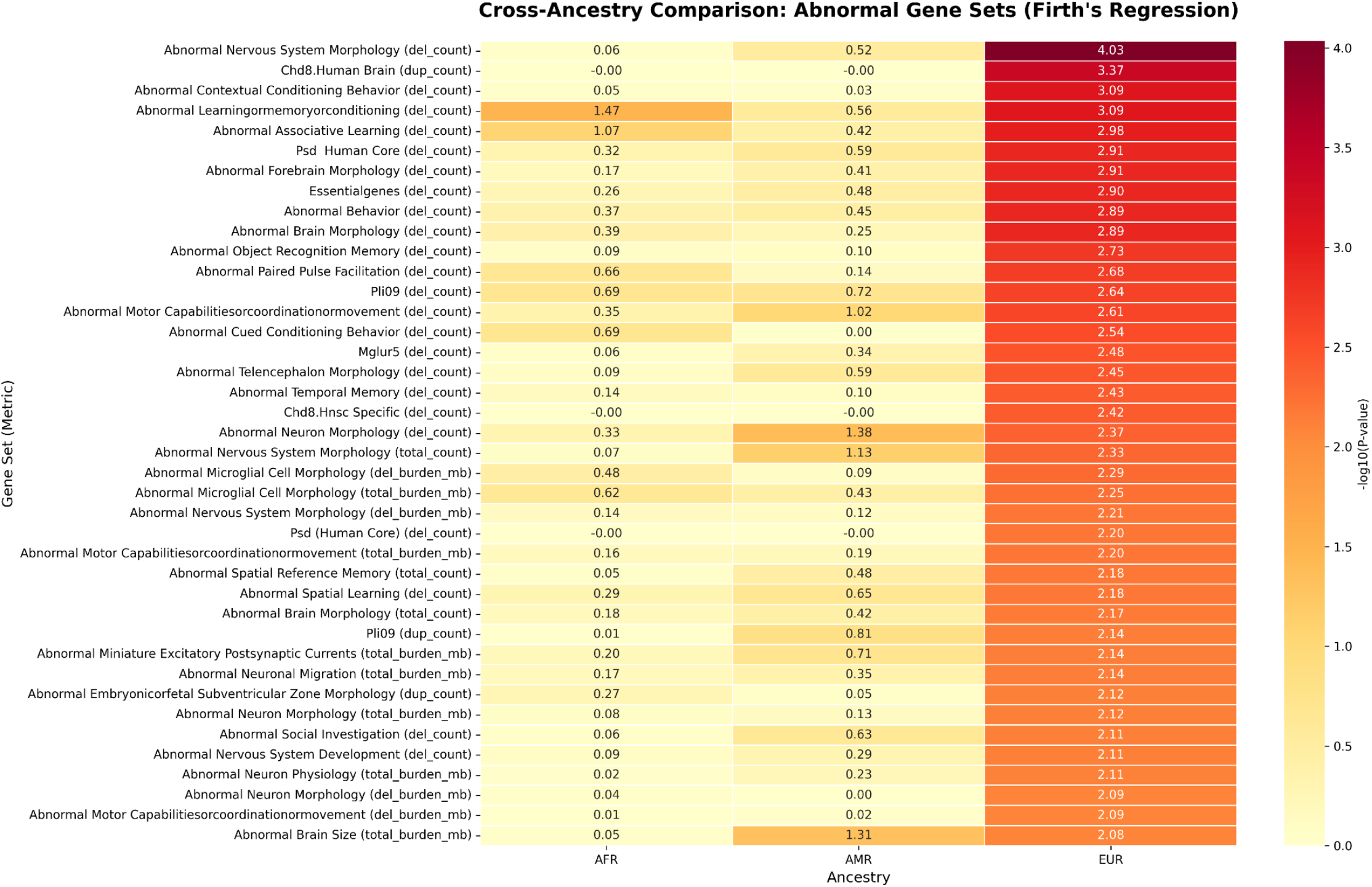
Multi-Ancestry Comparison of Gene Set Burden P-values.

**Supplementary Figure 20.**
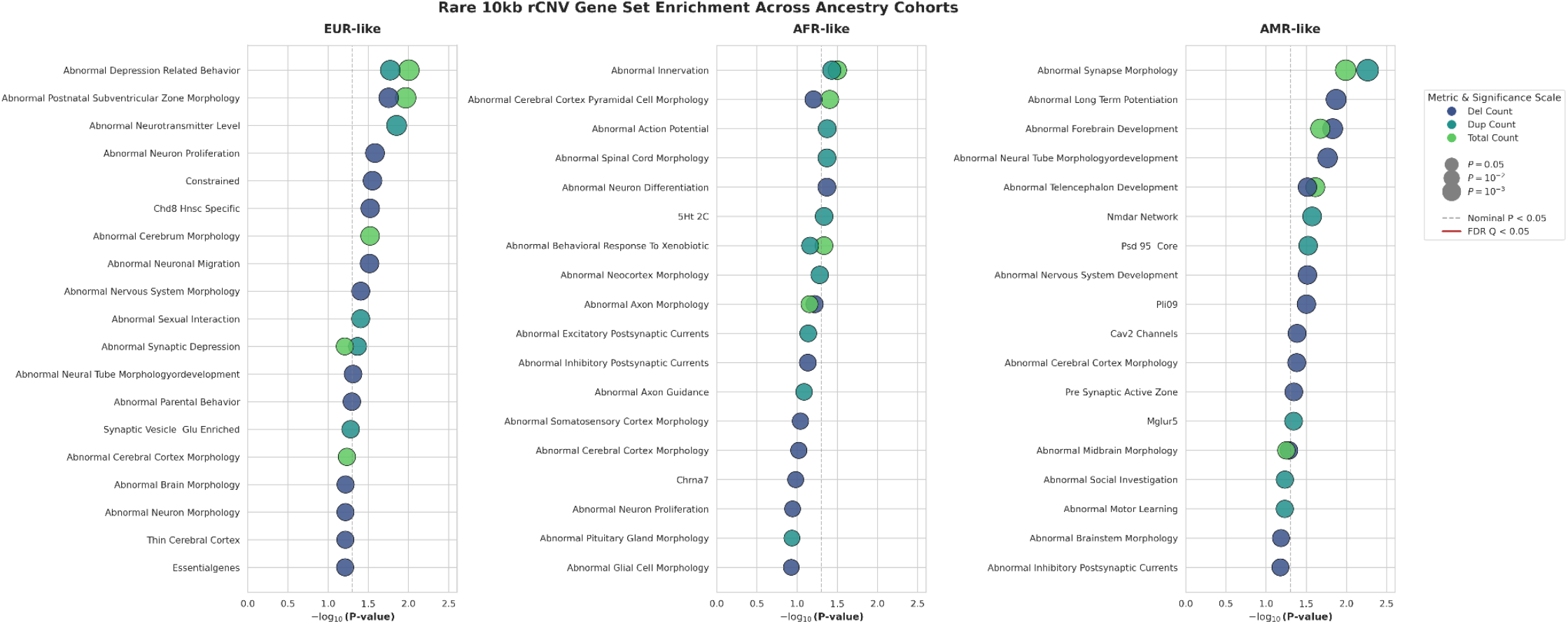
Cross ancestry comparison of abnormal mouse gene sets. It shows overall significant p-values in EUR-like ancestry group compared to AFR-like and AMR-like.

